# Pathogenesis-based pre-exposure prophylaxis associated with low risk of SARS-CoV-2 infection in healthcare workers at a designated Covid-19 hospital

**DOI:** 10.1101/2020.09.25.20199562

**Authors:** Michael V. Dubina, Veronika V. Gomonova, Anastasia E. Taraskina, Natalia V. Vasilyeva, Sergey A. Sayganov

**Author notes:** **Contact:** Dr. Dubina at the State Research Institute of Highly Pure Biopreparations FMBA Russia, 7 Pudozhskaya str., St. Petersburg 197110, Russia, or at.

## Abstract

At present, no agents are known to be effective in preventing Covid-19. Based on current knowledge of the pathogenesis of this disease, we suggest that SARS-CoV-2 infection might be attenuated by directly maintaining innate pulmonary redox, metabolic and dilation functions using well-tolerated medications that are known to serve these functions, specifically, using a low dose aerosolized combination of glutathione, inosine and potassium. From June 1 to July 10, 2020, we conducted a low-intervention open-label single-centre study to evaluate safety and efficacy of pre-exposure prophylaxis (PrEP) with the aerosolized combination medications (ACM) on SARS-CoV-2 incidence in 99 healthcare workers (HCWs) at a hospital that was designated to treat Covid-19 patients. We also retrospectively compared SARS-CoV-2 incidence in the ACM users to that in 268 untreated HCWs at the same hospital. Eligible participants received an aerosolized combination of 21.3 mg/ml glutathione, 8.7 mg/ml inosine in 107 mM potassium solution for 14 days. The main outcome was the frequency of laboratory confirmed SARS-CoV-2 cases, defined as individuals with positive genetic or immunological tests within 28 days of the study period. During the PrEP period, solicited adverse events occurred in five participants; all were mild and transient reactions. SARS-CoV-2 was detected in 2 ACM users (2%, 95% CI: 0.3% to 7.1%), which was significantly less than the incidence in 24 nonusers (9%, 95% CI: 5.8% to 13.0%; *P* = 0.02). Our findings might be used either to prevent SARS-CoV-2 infection, or to support ongoing and new research into more effective treatments for Covid-19. The study was registered with rosrid.ru, AAAA-A20-120061690058-2, and isrctn.com, ISRCTN34160010.

## INTRODUCTION

Covid-19, the novel infection caused by severe acute respiratory syndrome coronavirus 2 (SARS-CoV-2), was first detected in humans in December 2019 (*Zhu et al 2020*). As of September 20, 2020, more than 30 million confirmed cases and 950 thousand deaths have been reported worldwide (*WHO 2020 - ref*.*2*). The clinical hallmarks of the disease vary from asymptomatic infection to pneumonias featuring ground-glass opacities that might progress to life-threatening complications such as acute respiratory distress syndrome (ARDS) and multisystem organ failure (*Guan et al 2020*). Older patients and those with preexisting respiratory, cardiovascular and metabolic conditions appear to be at the greatest risk for severe complications and death (*Wu et al 2020*). A notable genomic feature of SARS-CoV-2 is its long spike (S) glycoprotein, with which has an approximately 80% overlapping sequence identity with those of previous SARS-CoV and MERS viruses (*Lu et al 2020*). Based on the similarities and past studies of SARS-CoVs, the pathogenesis of Covid-19 likely includes phases of SARS-CoV-2 infection that correspond to viral entry, replication in the upper airway and migration down the respiratory tract, with a robust immune response that triggers hypoxia and progression to ARDS (*Cao & Li 2020*). In the absence of a proven effective therapy, off-label or compassionate-use compounds have been used to treat Covid-19 with the assumption that airway epithelium damage, a diverse immune response and inflammation predominate in SARS-CoV-2 pathogenicity (*Baden & Rubin 2020*). Though there are currently more than one hundred Covid-19 candidate vaccines in development (*WHO 2020 - ref*.*8*), to date, no agents are known to be effective in preventing SARS-CoV-2 infection (*NIH 2020*).

Angiotensin-converting enzyme 2 (ACE2) has been identified to act as a receptor and an entry point for SARS-CoV-2, similar to what has been previously identified with SARS-CoVs (*Hoffmann et al 2020*). However, structural, biochemical and modeling data have revealed that SARS-CoV-2 might recognize ACE2 with a binding affinity that is an order of a magnitude greater than that for SARS-CoV (*Shang et al 2020, Wrapp et al 2020*). Marked ACE2 expression was found in the nose and alveolar epithelial cells, and presented in arterial smooth muscle cells and endothelial cells (*Hamming et al 2004, Hou et al 2020*), although there were found generally low ACE2 expression in cells from airways (*Sungnak et al 2020*). ACE2 represses the production of angiotensin II (Ang II) by angiotensin-converting enzyme (*Donoghue et al 2000*). Previous studies have reported considerably reduced ACE2 expression in the lungs of wild-type mice infected with SARS-CoV, suggesting that ACE2 might have a role in SARS-CoV-mediated severe acute lung pathologies (*Kuba et al 2005*). When SARS-CoVs interact with ACE2 to gain entry into cells, the downregulation of ACE2 – either directly due to viral binding or indirectly due to cell lysis – decreases Ang II inhibition (*Rivellese & Prediletto 2020*). Therefore, SARS-CoV-2 can reduce ACE2 activity and receptor consumption, further exacerbating an ACE2/Ang II regulatory imbalance (*Cheng et al 2020*), resulting in hypoxic pulmonary vasoconstriction (HPV) and impaired metabolic homeostasis in smooth muscle cells (SMCs) of distal bronchiolar airways.

Taken together, the key role of ACE2 in SARS-CoV-2 infection, the exceeding viral binding affinity of SARS-CoV-2 to the ACE2 receptor, and the documented severity of Covid-19 in subjects with respiratory, cardiovascular and metabolic conditions led us to hypothesize that the HPV and impaired airway SMC metabolic homeostasis, resulting from the robust SARS-CoV-2 virus-induced ACE2/Ang II regulatory imbalance, might prevail over the diverse immune response at the initial phase of Covid-19 pathogenesis. Though HPV is a physiological mechanism by which pulmonary arteries maintain blood oxygenation during alveolar hypoxia (*Sylvester et al 2012*), Ang II can induce robust pulmonary vasoconstriction via angiotensin receptor 1 (AT1R) (*Forrester et al 2018*), and causes internalization and degradation of membrane Ca2+-activated K+ channels (BKCa), providing an additional mechanism to contribute to vasoconstriction (*Leo et al 2015*). In addition, hypoxia, *per se*, might act either directly on peripheral pulmonary arterial SMCs either to inhibit a voltage-sensitive K^+^ channel (Kv) and induce membrane depolarization, and contraction (*Patel et al 1997*) or indirectly to stimulate release of vasoconstrictors and/or inhibit release of vasodilators (*Sato et al 2000*). Genome-wide transcriptome analyses *in vivo* after Ang II infusion have revealed upregulated genes in metabolism and ion transport pathways, while genes protective against oxidative stress, including glutathione synthetase and mitochondrial superoxide dismutase 2, were downregulated (*Makhanova et al 2010*). The disruption of reduction-oxidation (redox) signaling and excessive generation of reactive oxygen species by the injured pulmonary endothelium/epithelium under pathological conditions leads to increased endothelial permeability and upregulated expression of proinflammatory cytokines and adhesion molecules, amplifying tissue damage and pulmonary edema (*Kellner et al 2017*) Therefore, we hypothesized that SARS-CoV-2 infectivity and pathogenicity, and the resulting pulmonary abnormalities induced by an ACE2/Ang II regulatory imbalance, might be attenuated by directly maintaining the innate redox, dilation and metabolic functions of the lung using appropriate and well-tolerated medications.

There are currently available safe medications that have been previously reported to maintain ventilation-perfusion functions of the lung. A prime example is glutathione (γ-L-glutamyl-L-cysteinyl-glycine, GSH), a main nonprotein thiol (*Meister & Anderson 1983*), that decreases in alveolar epithelial cells upon exposure to toxins or respiratory viruses (*Mulier et al 1998*), and this decrease is associated with increased superoxide production and proinflammatory cytokine release (*Papi et al 2008*). It has been reported that GSH can be delivered by aerosol to directly augment the GSH level in the epithelial lining fluid of the lower respiratory tract *in vivo* (*Buhl et al 1990*) and to improve clinical outcome in patients with cystic fibrosis (*Bishop et al 2005*). GSH treatment of isolated bronchi *in vitro* resulted in decreases in SMCs contraction and bronchodilation (*Casoni et al 2003*), which increases membrane hyperpolarization via potassium (K+) channels on airway SMCs (*Deshpande et al 2010*). In the human lung, Kv and BKCa channels located in the apical membrane contribute to high K+ content in adult airway surface liquid (*Valeyre et al 1991*), and extracellular K+ is involved in matching tissue blood flow as a mediator of functional vasodilation (*Haddy et al 2006*). Moreover, vasodilators that act through receptors coupled to guanine nucleotide binding protein (G-protein) activate K+ channels through the cAMP signaling cascade, which includes adenosine (*Kleppisch & Nelson 1995*). The hydrolytic deamination of adenosine generates inosine, which increases in the extracellular space under metabolically stressful conditions and has been shown to have powerful immunomodulatory and cytoprotective activity by binding to G-protein-coupled A2A adenosine receptors (*Hasko et al 2008*). Inosine might serve as an alternative substrate for ATP generation during hypoxia (*Módis et al 2009*) and protects the bronchial and alveolar epithelium from the potentially deleterious consequences of neutrophil accumulation (*Qiu et al 2000*). Inosine might also exert antiviral effects through the incorporation into double-stranded viral RNA and potentiation of immune system sensing (*Sarvestani et al 2014*). It is important to note here that inosine-based compounds are amongst the drugs that being re-purposed for management of Covid-19 (*Kumar et al 2020*), and the efficacy of high-dose glutathione therapy in relieving dyspnea associated with Covid-19 pneumonia has been recently reported (*Horowitz et al 2020*).

Healthcare workers (HCWs) are at particularly high risk of acquiring SARS-CoV-2, from repeated exposure to infected patients (*Hunter et al 2020, Suárez-García et al 2020*). Based on the high risk of the infection in this population, the rationale presented above and the absence of effective agents to prevent or treat Covid-19, we aimed to evaluate whether pre-exposure prophylaxis (PrEP) using a low-dose aerosolized combination of glutathione, inosine and potassium might prevent SARS-CoV-2 acquisition in healthy adults who are at high risk for exposure to this infection.

## MATERIAL AND METHODS

### Study design and participants

A low-intervention prospective open-label investigator-initiated study was designed and conducted from June 1, 2020, to July 10, 2020. The study population was defined as healthy individuals who deliver care and services to Covid-19 patients (healthcare workers, HCWs), either directly as physicians or nurses or indirectly as assistants, technicians, or other support staff, using mandatory personal protective equipment in accordance with national guidelines. The study was initiated and conducted at North-Western State Medical University named after I.I. Mechnikov (NWSMU), a large healthcare center with 100 clinics and 4,500 employees. The study population was healthy voluntaries who were virus- and sera-negative from among the HCWs at a 264-bed hospital in NWSMU that was governmentally designated to treat Covid-19 patients as of May 5, 2020. Based on previous observational trials in high risk individuals exposed to routine Covid-19 positive contacts, a frequency of confirmed SARS-CoV-2 infection is not less than 11% (*Hunter et al 2020, Suárez-García et al 2020*). With 96 or more healthy volunteers in PrEP treatment group we expected significant decrease to 3% or less of SARS-CoV-2 positive cases during the study period. Full details of the study design and conduct can be seen in the protocol (Supplement 1). The Ministry of Health of the Russian Federation and the Federal Supervision Service for Healthcare provided oversight for the study in accordance with the framework of the Government Decree of the Russian Federation 441 of April 3, 2020. The study was reviewed and approved by the Local Ethics Committee at NWSMU (4/27-05-2020). Informed consent was obtained from all participants of the study. The data from the comparison group was collected in accordance with local routine Covid-19 surveillance reports. NWSMU collected the data and monitored the program. The study was registered with rosrid.ru, AAAA-A20-120061690058-2 and isrctn.com, ISRCTN34160010.

### Intervention

We used two solutions to prepare a low-dose aerosolized combination of three medications; the two solutions were 3% inosine-glutamyl-cysteinyl-glycine disodium (i.e., inosine-glutathione; Molixan, PharmaVAM; St. Petersburg, Russia) – a patented metabolic agent that is approved in Russia for parenteral use in combination treatment of viral hepatitis (grls.rosminzdrav.ru, N001355/02), and 4% potassium chloride (Solopharm, St. Petersburg, Russia). The medication for each 5-minute inhalation session was prepared *ex tempore* by mixing solutions of 1.0 ml inosine-glutathione and 0.25 ml potassium chloride to achieve the final concentrations: 21.3 mg/ml glutathione, 8.7 mg/ml inosine and 107 mM potassium (pH 5.5). The combined medication was self-administered as an aerosol using a personal handheld nebulizer (Nebzmart MBPN002; MicroBase Technology, Taiwan) driven by compressed air at 0.25 ml/min. Eligible participants received the study treatment for 14 days; they were instructed to perform four inhalation sessions per day with 4 h in between sessions.

### Study procedures

From the full staff of HCWs at the hospital, 99 volunteers were assigned to a study condition among 100 HCWs who were randomly recruited to participate in the study by members of the study team at the hospital from June 1 to June 12, 2020 (Figure 1). All participants completed a pre-enrollment evaluation, which included data from genetic and serology SARS-CoV-2 tests performed prior to enrollment. Data on treatment adherence and adverse events were collected on day 7 and day 14; additional follow-up information was solicited through day 28. The primary outcome was new-onset SARS-CoV-2 infection, detected by genetic or immunological tests over the study period. A comparison group of HCWs from the hospital was analyzed retrospectively among HCWs who were not included in the study group, were virus- and sera-negative at the end of May, 2020 and either worked until the final date of the study. All HCWs in the hospital were tested for SARS-CoV-2 weekly in accordance with national guidelines and recommendations. Additionally, all HCWs received qualitative serological testing for IgM and IgG against SARS-CoV-2 at the end of May and beginning of July, 2020. Therefore, outcome data in the comparison group were collected from the results of these routine tests.

**Figure 1.**
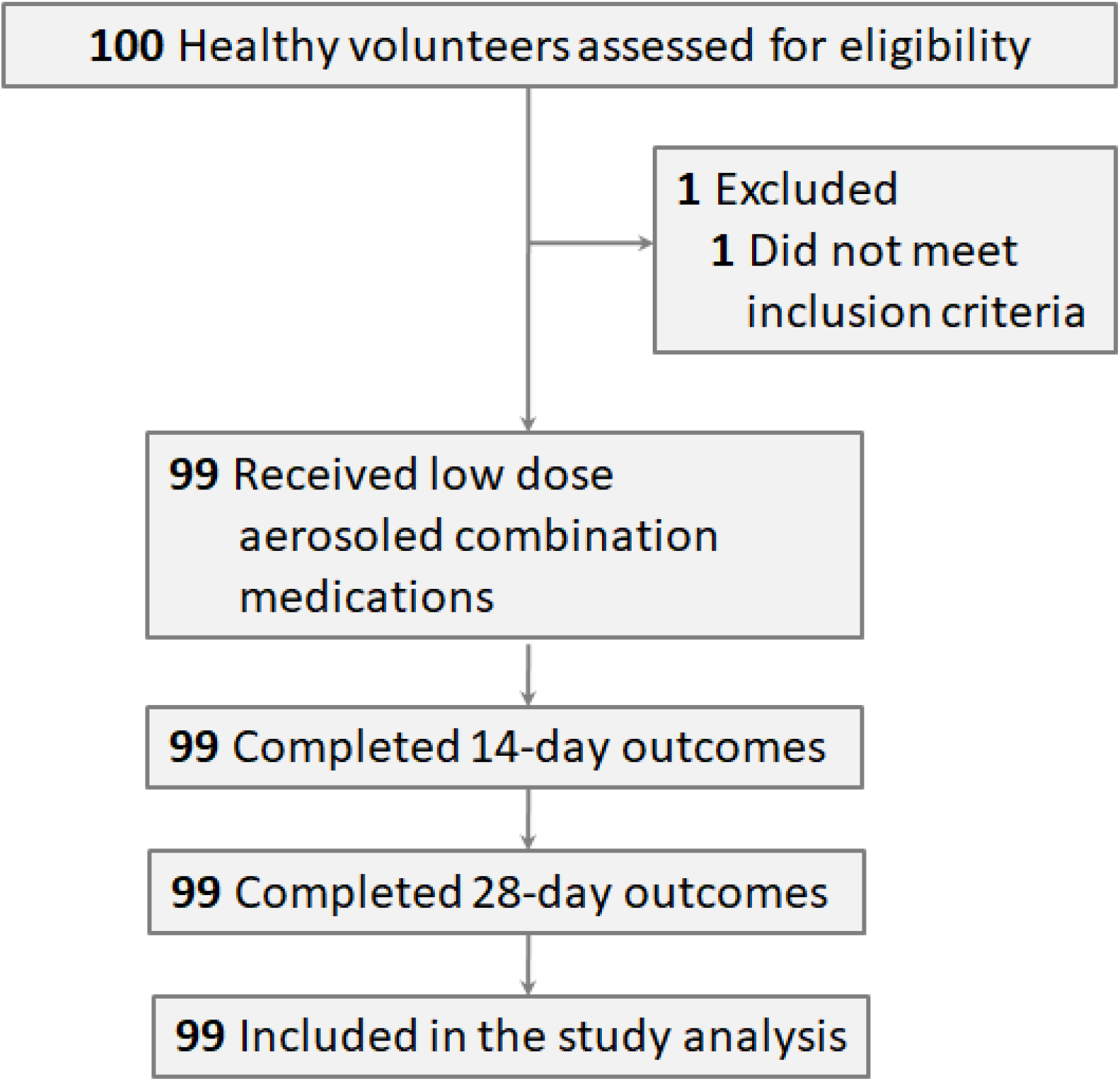
Flow diagram showing the progress of healthy subjects throughout the study.

### SARS-CoV-2 testing

Nasopharyngeal swabs were collected for genetic detection of SARS-CoV-2 by real-time polymerase chain reaction (RT-PCR), and venous blood draws were performed for immunological assessments. All samples were de-identified, transported to the NWSMU diagnostic laboratory within 3h, and stored at +4°C until analysis. Total RNA was extracted from the swabs using an RNA-Express Kit (Lytech, Moscow, Russia). SARS-CoV-2 was tested by RT-PCR using two target genes, namely, the RNA-dependent RNA polymerase (RdRp) gene of the open reading frame1ab (ORF1ab) sequence and envelope (E) gene, with the POLIVIR SARS-CoV-2 Assay (Lytech, Moscow, Russia) using the CFX-96 Touch system (Bio-Rad, Hercules, USA). A result was considered positive if the cycle threshold was below 35 for both target genes. Serum antibodies were assayed using two diagnostic kits approved in Russia for qualitative detection of immunoglobulins against SARS-CoV-2 in human plasma, per the manufacturers’ instructions: SARS-CoV-2-IgM-IFA BEST (Vector-Best, Novosibirsk, Russia) and SARS-CoV-2–IgG (Lytech, Moscow, Russia). The antibodies were measured on a HumaReader HS microtiter plate reader (HUMAN, Wiesbaden, Germany) with spectrophotometric method of optical density at 450 nm.

### Statistical analysis

The chi-square test was used to compare theSARS-CoV-2 incidence between the study group and the comparison group; p-value less than 0.05 was considered statistically significant. Because the analysis did not include a provision for correcting for multiple comparisons in the tests for association between baseline variables and outcomes, the results are reported as point estimates and 95% confidence intervals. The analysis was conducted using SAS software.

## RESULTS

During the study period (June 1, 2020, to July 10, 2020), the cumulative number of patients with Covid-19 in the hospital increased from 447 to 884 (97.8%). Among the 410 HCWs who worked at the hospital, 43 (10.5%) were SARS-CoV-2 positive before the beginning of the study (n=29), or did not work until the final date of the study (n=14); 99 HCWs were enrolled as participants in the study (24.1%); the remaining 268 HCWs were designated as the comparison group (65.4%).

The mean age of the participants was 27.0 years (95% CI: 25.3 to 28.7), 69% were female, and 52% were nurses (51/99). The demographic characteristics of the participants and the comparison group did not differ significantly (Table 1).

**Table 1.** Baseline characteristics of HCWs in the Covid-19 hopsital.*

| Characteristic | ACM users (n=99) | Nonusers (n=268) |
| --- | --- | --- |
| Demographic |  |  |
| Median age – yrs (IQR) | 25 (5) | 25 (11) |
| Age category – No. (%) |  |  |
| < 40 | 88 (89) | 228 (85) |
| 40 to 65 yrs | 11 (11) | 40 (15) |
| Male sex –No. (%) | 31 (31) | 106 (40) |
| Professional |  |  |
| Physicians – No. (%) | 38 (38) | 99 (37) |
| Nurses – No. (%) | 51 (52) | 140 (52) |
| Others – No. (%) | 10 (10) | 29 (11) |

\* HCWs – healthcare workers. IQR – interquartile range. ACM – aerosoled combination medication.

During the study period, no serious adverse events were observed in the ACM group, and none of the prespecified stopping rules were met. During the treatment period, solicited systemic and common adverse events, which occurred in five participants (5%), included headache in two participants (2%), itchy throat in two participants (2%) and dry cough in one participant (1%); all were mild and transient reactions lasting no longer than 30 min during or after the inhalation session. The adverse events revealed no patterns of concern; neither treatment interruption nor modification was indicated.

The total incidence of SARS-CoV-2 detected in the ACM group was 2% (2/99; 95% CI: 0.3% to 7.1%), which was significantly less than the virus- or sera-positivity rate in the comparison group, which was 9% (24/268; 95% CI: 5.8% to 13.0%; *P* = 0.02) (Figure 2).

**Figure 2.**
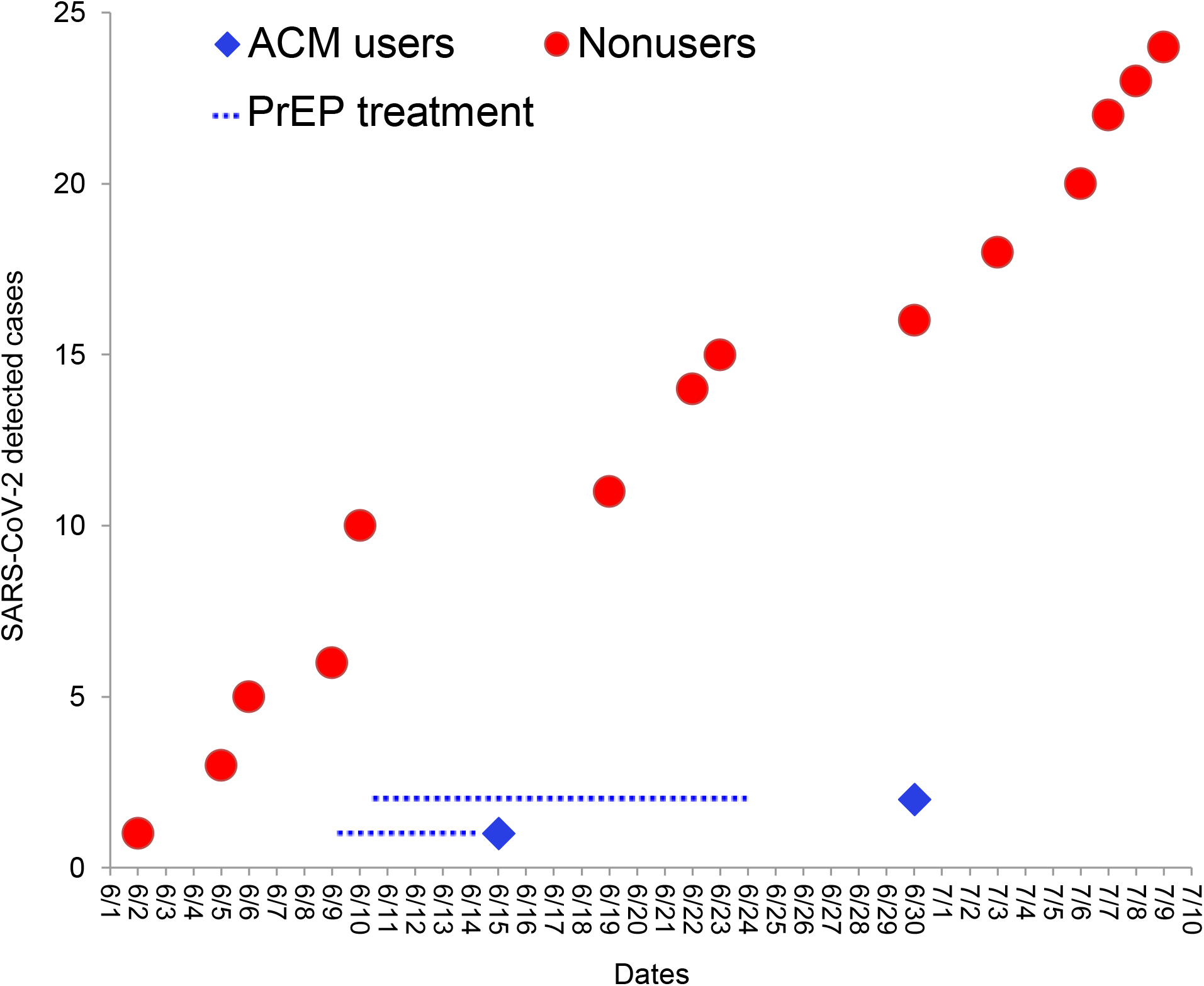
Cumulative SARS-CoV-2 incidence in HCWs*. *HCWs – healthcare workers. ACM – aerosoled combination medication. PrEP – pre-exposure prophylaxis with ACM.

## DISCUSSION

Here, we report that the pathogenesis-based prophylaxis associated with significant reduction of SARS-CoV-2 incidence to 2% in HCWs who delivered care and services to patients with Covid-19 compared with 9% incidence in remaining HCWs who did not use the treatment at the same hospital during the entire study period. Effective PrEP for Covid-19 would not only safely prevent high-risk individuals from acquiring SARS-CoV-2 infection from close contacts infected with the virus but also could attenuate the infection if it did occur. At present, most clinical trials of pharmacologic interventions to prevent SARS-CoV-2 infection have been conducted with medications that were previously used in the treatment of Covid-19 (e.g., hydroxychloroquine), and have focused on evaluating the safety of these drugs in this new context (*Cohen 2020*). To date, this repurposing of established drugs and the development of vaccines has been mainly driven by the common approach of considering SARS-CoV-2 virulence and the host immune reaction as the primary factors in the pathogenesis of Covid-19 (*Cao & Li 2020*). In contrast, based on the triggering role of an ACE2/Ang II imbalance in SARS-CoV-2 pathogenicity (*Cheng et al 2020, Rivellese & Prediletto 2020*), we hypothesized that Covid-19 might be attenuated by direct maintenance of innate pulmonary redox, dilation and metabolic functions using a combination of medications that were previously reported to be well tolerated after aerosol delivery to human airways. Using an aerosolized combination of glutathione, potassium and inosine at low doses, we designed and conducted a low-intervention prospective open-label study of PrEP in healthy participants at high risk of acquiring SARS-CoV-2 due to their repeated exposure to confirmed Covid-19 patients. Here we have shown the safety and efficacy of the pathogenesis-based PrEP for SARS-CoV-2 infection, and also highlight the following aspects in address of Covid-19 prevention and treatment.

In the study, ACM had no serious adverse events at the chosen dose of 21.3 mg ml/1 glutathione and 8.7 mg ml/1 inosine in hypo-osmolar potassium chloride solution. This is consistent with previous data that showed good tolerance of the inhaled components of ACM separately at doses up to an order of magnitude greater in healthy participants or patients. In particular, no toxic or adverse effects were observed for inhaled glutathione at doses of 300 mg or more when administered to participants with pulmonary fibrosis (*Griese et al 2004*). Inosine solution, when inhaled at concentrations up to 373 µmol (100 mg)/ml, had no effect on airway responses in healthy and asthmatic participants (*Mann et al 1986*). Therefore, a combination of these medications at low doses, used simultaneously, might be an optimal way to attenuate the main steps of Covid-19 pathogenesis. Importantly, some of the agents can exert a number of physiological effects, increasing the cumulative antiviral efficiency of the combined medication. For instance, in addition to the metabolic activity of inosine, adenosine to inosine modification (A-to-I) by adenosine deaminases acting on RNA is a recently discovered process of posttranscriptional pre-mRNA modification (*Nishikura 2016*). This process exerts antiviral effects through the incorporation of inosine into double-stranded viral RNA followed by RNase-specific viral degradation (*Scadden & Smith 1997*) and through potentiation of immune system sensing (*Sarvestani et al 2014*).

The significant reduction of SARS-CoV-2 incidence among HCWs treated with ACM in our study might support the hypothesized importance of the virus-induced ACE2/Ang II regulatory imbalance in human airways, along with the consequent robust pulmonary hypoxia and peripheral ventilation-perfusion abnormalities. Moreover, these results suggest the important prevalence of these processes over the diverse host immune response and viral load, *per se*, in the pathogenesis of Covid-19. Our approach and findings might be used either to prevent SARS-CoV-2 infection or, even more effectively, to support ongoing and new research into more effective treatments for Covid-19. Further studies of pathogenesis-based combination medications for the prophylaxis and treatment of this severe disease need to be performed in greater detail using multi-centre randomized placebo-controlled methods.

## Supporting information

Protocol

TREND Statement Checklist

## Data Availability

The datasets generated during and/or analysed during the study are available from the corresponding author on reasonable request.

This study was funded by North-Western State Medical University named after I.I. Mechnikov (NWSMU) from personal charity financing.

All authors declare no conflict of interest. NWSMU made all decisions regarding study design and implementation. PharmaVAM donated medicinal products for use in this study but did not provide any financial support.

The first draft of the manuscript was written by the first author with contributions from all authors, and the last author as the senior author. All authors had full access to raw data, and assume responsibility for accuracy and completeness of the reported data.

The views expressed in this manuscript are those of Dr. Dubina. The conclusions, findings, and opinions expressed by authors contributing to this manuscript does not necessarily reflect the official position or policies of any governmental authorities. Any mention of trade names, commercial products or organizations imply endorsement by any of the groups mentioned above.

We thank all the participants for their altruism and their dedication to this study, and Vladimir S. Litvinenko for his invaluable support. We thank Otari G. Khurzilava, DMSc, at NWSMU for his senior mentorship; Anna M. Bitieva, MD; Anastasia S. Fedorenko, MD, PhD; Tatyana S. Fil, MD, PhD; Olga V. Kovaleva, MD; Natalia A. Prokofeva, MD; Sergey V. Tikhonov, MD, PhD; Natalia A. Trostianetckaia, MD, PhD, and Daredjan B. Tcurtcumiya, MD, PhD, as the members of the study team at NWSMU for their contributions; Yulia V. Borzova, PhD, and Olga A. Shurpitskaya at Kashkin Institute of Medical Micology of NWSMU for providing the immunologic assays; Elgudga L. Lataria, MD, PhD; Elena I. Mazenko; Elena E. Shaduyko, and Alexey V. Silin, DMSc, at NWSMU for organizational support; Alexei A. Bogdanov, PhD, at the State Research Institute of Highly Pure Biopreparations (SRIHPB) for providing statistical analysis and technical assistance. Dr. Dubina is personally thankful to Ekaterina Allora; Fedor V. Moiseyenko, DMSc; Eugene Sokolov; Vera V. Vysochinskaya MD; Nikita Zaguskin; Irina B. Zueva, DMSc, and others for their valuable advice and genuine encouragement.

## REFERENCES

1. Zhu N, Zhang D, Wang W, et al. A novel coronavirus from patients with pneumonia in China, 2019. N Engl J Med 2020;382:727–733.

2. World Health Organization. Novel coronavirus (2019-nCoV) situation reports. WHO September 21, 2020. (http://www.who.int/emergencies/diseases/novel-coronavirus-2019/situation-reports/)

3. Guan WJ, Ni ZY, Hu Y, et al. Clinical characteristics of coronavirus disease 2019 in China. N Engl J Med 2020;382:1708–1720.

4. Wu C, Chen X, Cai Y, et al. Risk factors associated with acute respiratory distress syndrome and death in patients with coronavirus disease 2019 pneumonia in Wuhan, China. JAMA Intern Med 2020;180:1–11.

5. Lu R, Zhao X, Li J, et al. Genomic characterization and epidemiology of 2019 novel coronavirus: implications for virus origins and receptor binding. Lancet 2020;395:565–574.

6. Cao W, Li T. COVID-19: towards understanding of pathogenesis. Cell Research 2020;30:367–369

7. Baden LR, Rubin EJ. Covid-19 – the search for effective therapy. N Engl J Med 2020;382:1851–1852.

8. World Health Organization. Draft landscape of COVID-19 candidate vaccines. WHO September 21, 2020. (https://www.who.int/who-documents-detail/draft-landscape-of-covid-19-candidate-vaccines)

9. National Institutes of Health. Coronavirus disease 2019 (COVID-19) treatment guidelines. NIH April 21, 2020. (https://www.covid19treatmentguidelines.nih.gov/overview/prophylaxis)

10. Hoffmann M, Kleine-Weber H, Schroeder S, et al. SARS-CoV-2 cell entry depends on ACE2 and TMPRSS2 and is blocked by a clinically proven protease inhibitor. Cell 2020;181:271–280.

11. Shang J, Ye G, Shi K, et al. Structural basis of receptor recognition by SARS-CoV-2. Nature 2020;581:221–224.

12. Wrapp D, Wang N, Corbett KS, et al. Cryo-EM structure of the 2019-nCoV spike in the prefusion conformation. Science 2020;367:1260.

13. Hamming I, Timens W, Bulthuis M, Lely A, Navis G, and Van Goor H. Tissue distribution of ACE2 protein, the functional receptor for SARS coronavirus. A first step in understanding SARS pathogenesis. J Pathol 2004;203:631–637.

14. Hou YJ, Okuda K, Edwards CE, et al. SARS-CoV-2 Reverse genetics reveals a variable infection gradient in the respiratory tract. Cell 2020;182:1–18.

15. Sungnak W, Huang N, Bécavin C, et al. SARS-CoV-2 entry factors are highly expressed in nasal epithelial cells together with innate immune genes. Nat Med 2020;26:681–687.

16. Donoghue M, Hsieh F, Baronas E, et al. A novel angiotensin-converting enzyme-related carboxypeptidase (ACE2) converts angiotensin I to angiotensin 1-9. Circ Res 2000;87:E1–E9.

17. Kuba K, Imai Y, Rao S, et al. A crucial role of angiotensin converting enzyme 2 (ACE2) in SARS coronavirus–induced lung injury. Nat Med 2005;11:875–879.

18. Rivellese F, Prediletto E. ACE2 at the centre of COVID-19 from paucisymptomatic infections to severe pneumonia. Autoimmun Rev 2020;19:102536.

19. Cheng H, Wang Y, and Wang G-Q. Organ-protective effect of angiotensin-converting enzyme 2 and its effect on the prognosis of COVID-19. J Med Virol 2020;92:726–730.

20. Sylvester JT, Shimoda LA, Aaronson PI, and Ward JP. Hypoxic pulmonary vasoconstriction. Physiol Rev 2012;92:367–520.

21. Forrester SJ, Booz GW, Sigmund CD, et al. Angiotensin II signal transduction: an update on mechanisms of physiology and pathophysiology. Physiol Rev 2018;98:1627–1738.

22. Leo MD, Bulley S, Bannister JP, Kuruvilla KP, Narayanan D, and Jaggar JH. Angiotensin II stimulates internalization and degradation of arterial myocyte plasma membrane BK channels to induce vasoconstriction. Am J Physiol Cell Physiol 2015;309:C392–C402.

23. Patel AJ, Lazdunski M, and Honore E. Kv2.1/Kv9, a novel ATP-dependent delayed-rectifier K+ channel in oxygen-sensitive pulmonary artery myocytes. EMBO J 1997;16:6615–6625.

24. Sato K, Morio Y, Morris KG, Rodman DM, and McMurtry IF. Mechanism of hypoxic pulmonary vasoconstriction involves ET(A) receptor-mediated inhibition of K(ATP) channel. Am J Physiol Lung Cell Mol Physiol 2000;278:L434–L442.

25. Makhanova NA, Crowley SD, Griffiths RC, and Coffman TM. Gene expression profiles linked to AT1 angiotensin receptors in the kidney. Physiol Genomics 2010;42A:211–218.

26. Kellner M, Noonepalle S, Lu Q, Srivastava A, Zemskov E, and Black SM. R OS signaling in the pathogenesis of acute lung Injury (ALI) and acute respiratory distress syndrome (ARDS). Adv Exp Med Biol 2017;967:105–137.

27. Meister A, Anderson ME. Glutathione. Annu Rev Biochem 1983;52:711–760.

28. Mulier B, Rahman I, Watchorn T, Donaldson K, MacNee W, and Jeffery PK. Hydrogen peroxide-induced epithelial injury: the protective role of intracellular nonprotein thiols (NPSH). Eur Respir J 1998;11:384–391.

29. Papi A, Contoli M, Gasparini P, et al. Role of xanthine oxidase activation and reduced glutathione depletion in rhinovirus induction of inflammation in respiratory epithelial cells. J Biol Chem 2008;283:28595–28606.

30. Buhl R, Vogelmeier C, Critenden M, et al. Augmentation of glutathione in the fluid lining the epithelium of the lower respiratory tract by directly administering glutathione aerosol. PNAS 1990;87:4063–4067.

31. Bishop C, Hudson VM, Hilton SC, and Wilde C. A pilot study of the effect of inhaled buffered reduced glutathione on the clinical status of patients with cystic fibrosis. Chest 2005;127:308–317.

32. Casoni GL, Chitano P, Pinamonti S, et al. Reducing agents inhibit the contractile response of isolated guinea-pig main bronchi. Clin Exp Allergy 2003;33:999–1004.

33. Deshpande DA, Wang WC, McIlmoyle EL, et al. Bitter taste receptors on airway smooth muscle bronchodilate by localized calcium signaling and reverse obstruction. Nat Med 2010;16:1299–1304.

34. Valeyre D, Soler P, Basset G, et al. Glucose, K+, and albumin concentrations in the alveolar milieu of normal humans and pulmonary sarcoidosis patients. Am Rev Respir Dis 1991;143:1096–1101.

35. Haddy FJ, Vanhoutte PM, and Feletou M. Role of potassium in regulating blood flow and blood pressure. Am J Physiol Regul Integr Comp Physiol 2006;290:R546–R552.

36. Kleppisch T, Nelson MT. Adenosine activates ATP-sensitive potassium channels in arterial myocytes via A2 receptors and cAMP-dependent protein kinase. PNAS 1995;92:12441-12445.

37. Hasko G, Linden J, Cronstein B, and Pacher P. Adenosine receptors: therapeutic aspects for inflammatory and immune diseases. Nat Rev Drug Discov 2008;7:759–770.

38. Módis K, Gero D, Nagy N, Szoleczky P, Tóth ZD, and Szabó C. Cytoprotective effects of adenosine and inosine in an in vitro model of acute tubular necrosis. Br J Pharmacol 2009;158:1565–1578.

39. Qiu FH, Wada K, Stahl GL, and Serhan CN. IMP and AMP deaminase in reperfusion injury down-regulates neutrophil recruitment. PNAS 2000;97:4267–4272.

40. Sarvestani ST, Tate MD, Moffat JM, et al. Inosine-mediated modulation of RNA sensing by Toll-like receptor 7 (TLR7) and TLR8. J Virol 2014;88:799–810.

41. Kumar R, Gupta N, Kodan P, Mittal A, Soneja M, and Wig N. Battling COVID-19: using old weapons for a new enemy. Trop Dis Travel Med Vaccines 2020; 6.

42. Horowitz RI, Freeman PR, and Bruzzese J. Efficacy of glutathione therapy in relieving dyspnea associated with COVID-19 pneumonia: A report of 2 cases. Respir Med Case Rep 2020;30:101063.

43. Hunter E, Price DA, Murphy E, et al. First experience of COVID-19 screening of health-care workers in England. Lancet 2020;395:e77–e78.

44. Suárez-García I, Lobo Abascal P, Martínez de Aramayona López MJ, and Sáez Vicente A. SARS-CoV-2 infection among healthcare workers in a hospital in Madrid, Spain. J Hosp Infect 2020;106(2):357–363.

45. Cohen MS. Hydroxychloroquine for the prevention of Covid-19 - searching for evidence. N Engl J Med 2020;383:585–586.

46. Griese M, Ramakers J, Krasselt A, et al. Improvement of alveolar glutathione and lung function but not oxidative state in cystic fibrosis. Am J Respir Crit Care Med 2004;169:8 22–828.

47. Mann JS, Holgate ST, Renwick AG, and Cushley MJ. Airway effects of purine nucleosides and nucleotides and release with bronchial provocation in asthma. J Appl Physiol 1986;61:1667–1676.

48. Nishikura K. A-to-I editing of coding and non-coding RNAs by ADARs. Nat Rev Mol Cell Biol 2016;17:83–96.

49. Scadden AD, Smith CW. A ribonuclease specific for inosine-containing RNA: a potential role in antiviral defence? EMBO J 1997;16:2140–2149.

