## Supplementary material for "Pathogenesis-based pre-exposure prophylaxis associated with low risk of SARS-CoV-2 infection in healthcare workers at a designated Covid-19 hospital": Protocol

### MIBVD-19
### Low-intervention Study of the Aerosol Use of a Combination Medication as Pre-Exposure Prophylaxis for the Novel Coronavirus Infection COVID-19

#### Protocol
#### (Healthy Volunteer XXX)

Michael V. Dubina, MD, PhD, DSc (Med)
Academician of the Russian Academy of Sciences
19 Zamshina street 55, Saint-Petersburg
195221 Russian Federation

Version: 22 April 2020

**CONFIDENTIAL**

### TABLE OF CONTENTS

TABLE OF CONTENTS ... 2

1. BACKGROUND ... 3
   - 1.1. General Information ... 3
   - 1.2. Rationale for Use of Combination Medication ... 4
2. STUDY ASSESSMENT ... 9
   - 2.1. Objective ... 9
   - 2.2. Endpoints ... 9
   - 2.3. Design ... 9
   - 2.4. Population ... 9
   - 2.5. Inclusion Criteria ... 9
   - 2.6. Exclusion Criteria ... 9
   - 2.7. Early Discontinuation Criteria ... 10
   - 2.8. Duration ... 11
3. DESCRIPTION OF COMBINATION MEDICATION ... 12
4. DOSAGE AND ADMINISTRATION OF COMBINATION MEDICATION ... 14
5. CLINICAL AND LABORATORY ASSESSMENT ... 15
6. BENEFIT-RISK ASSESSMENT ... 16
7. STATISTICAL ANALYSIS ... 19
8. REFERENCES ... 20
9. APPENDICES ... 22
   - Appendix 1. Study Plan ... 22
   - Appendix 2. Informed Consent Form ... 23
   - Appendix 3. Instruction for the Use of Combination Medication ... 24
   - Appendix 4. Individual Record Form ... 25

### 1. BACKGROUND

#### 1.1. General Information

**Study title**

Low-interventional study of the aerosol combination therapy for pre-exposure prophylaxis of the novel coronavirus infection COVID-19

**Study type**

Low-interventional study (Order of the Government of the Russian Federation of April 3, 2020, No. 441 "On the aspects of the circulation of drugs for medical use intended for application in the conditions of risk, occurrence, and liquidation of an emergency situation and for the organization of medical assistance to persons affected by the emergency, prevention of emergency situations, prevention and treatment of the diseases that pose a threat to public health, diseases and injuries resulting from adverse chemical, biological, and radiation factors").

**Study code**

MIBVD-19

**Study phase**

Not applicable. Pilot study.

**Ethical considerations**

The study will be conducted in accordance with the Protocol and the legislation currently in force in the Russian Federation and is regulated by the following normative documents, as well as the available addendums and explanations to these documents:

- The Constitution of the Russian Federation;
- Federal Law of April 12, 2010, No. 61-FZ "On Circulation of Medicines";
- Federal Law of November 21, 2011, No. 323-FZ “On the Fundamentals of Public Health Protection in the Russian Federation”;
- National Standard of the Russian Federation GOST R52379-2005 “Good Clinical Practice”;
- Order of the Government of the Russian Federation of April 3, 2020, No. 441 "On the aspects of the circulation of drugs for medical use intended for application in the conditions of risk, occurrence, and liquidation of an emergency situation and for the organization of medical assistance to persons affected by the emergency, prevention of emergency situations, prevention and treatment of the diseases that pose a threat to public health, diseases and injuries resulting from of adverse chemical, biological, and radiation factors";
- Temporary guidelines of the Ministry of Health of the Russian Federation on the prevention, diagnosis, and treatment of the novel coronavirus infection (COVID-19) in the current edition;
- Federal Law of July 27, 2006, No. 152-FZ "On personal information";
- Order of the Ministry of Health of the Russian Federation of November 29, 2012, No. 986n “On approval of the Regulation of the Ethics Council”;

- Order of the Ministry of Health of the Russian Federation of December 27, 2012, No. 1570 “On the Composition of the Ethics Council”;
- Order of the Ministry of Health of the Russian Federation of April 1, 2016, No. 200n “On approval of the Rules of Good Clinical Practice";
- Eurasian Economic Commission Council Decision dated November 3, 2016, No. 79 "On approval of the Rules of Good Clinical Practice of the Eurasian Economic Union";
- Helsinki Declaration in its current version (Fortaleza, 2013).

#### 1.2. Rationale for Use of Combination Medication

**In December 2019**, an outbreak of a novel coronavirus infection was identified in the People's Republic of China (PRC) with an epicenter in Wuhan (Hubei province). A previously unknown single-stranded RNA virus from the Coronaviridae family and the β-CoV genus was detected by sequencing in samples obtained from patients with pneumonia (ZhuN. et al., 2020). On February 11, 2020, the International Committee on Taxonomy of Viruses designated the official name to the pathogen SARS-CoV-2. The World Health Organization (WHO) on February 11, 2020, announced the official name of the disease caused by the novel coronavirus COVID-19 ("Coronavirus disease 2019"). SARS-CoV-2 is included in the list of diseases that pose a threat to public health (Order of the Government of the Russian Federation of January 31, 2020, No. 66). Many aspects of coronavirus infection pathogenesis require further comprehensive study. Currently, there are no proven effective clinical protocols for the prevention and treatment of COVID-19 infection.

**Members of the coronavirus family** were previously regarded as agents present in the structure of acute respiratory viral infections causing mild or moderate upper respiratory tract disorders (with extremely rare lethal outcomes). At the end of 2002, SARS-CoV coronavirus emerged as the etiological agent of atypical pneumonia that caused severe acute respiratory syndrome (SARS) in humans. Overall, during the 2002–2004 epidemic, more than 8000 cases in 37 countries were registered, of which 774 were fatal. A new MERS-CoV coronavirus, the causal agent of Middle East respiratory syndrome, emerged in 2012. Until January 31, 2020, 2519 cases of coronavirus infection caused by this virus were registered, 866 of which were fatal (82% of cases were reported in Saudi Arabia).

**The source of COVID-19 infection** is a sick person, including one who is in the incubation period of the disease. The infection is transmitted through respiratory droplets, airborne transmission, and contact routes. The entrance pathway of SARS-CoV-2 is the epithelium of the upper respiratory tract, stomach and intestines. The initial stage of infection is the penetration of the pathogen into target cells (mainly pulmonary alveolar type II cells) that have angiotensin-converting enzyme 2 (ACE2) receptors. SARS-CoV-2 can disseminate from the systemic circulation into other organs or across the cribriform plate of the ethmoid bone to the CNS. COVID-19 is confirmed if laboratory testing of biological samples for the presence of SARS-CoV-2 RNA using nucleic acid amplification techniques gives a positive result, regardless of clinical symptoms. The incubation period for COVID-19 infection lasts between 2 and 14 days, with an average of 5–7 days.

**The initial clinical manifestations** are similar to those of other acute respiratory infections (body temperature above 37.5°C and one or more of the following symptoms: dry cough or cough with sparse sputum; shortness of breath; feeling of congestion in the chest; oxygen saturation of blood according to pulse oximetry ($SpO_2$) $\leq$ 95%; sore throat, runny nose, and other catarrhal symptoms; weakness; headache; anosmia; diarrhea) in the absence of other known reasons that explain the clinical picture, regardless of the epidemiological history. The early symptoms may include myalgia (11%), mental confusion (9%), headaches (8%), hemoptysis (5%), diarrhea (3%), nausea, vomiting, and heart palpitations. At the onset of infection, these symptoms can also be observed in the absence of fever. Eighty percent of patients experience a mild form of acute respiratory viral infection, and 20% of confirmed cases are severe.

**The most common complication** of COVID-19 is bilateral pneumonia with the following typical manifestations observed on computed tomography or chest radiography images of the lungs: a decrease in pneumatization of the lung tissue manifested as ground-glass opacity with or without consolidation and with or without thickening of the interlobular septa ("cobblestone" appearance); multiple sites of decreased pneumatization of lung tissue manifested as a rounded shape ground-glass opacity with or without consolidation and with or without thickening of the septa ("cobblestone" appearance); a reverse halo or other signs of organizing pneumonia (at the later disease stages); predominantly peripheral localization; and bilateral nature of the disorder. Radiological signs of inflammatory lesions on computed tomography images may be absent in 18% of patients with a mild condition and in 3% of patients in a moderate/severe state, as well as in the early stages of the disease.

**Severe forms** of COVID-19 develop mainly in elderly patients (65 years or older). Among patients, concomitant diseases are often observed: diabetes mellitus (20%), arterial hypertension (15%), and other cardiovascular diseases (15%). In severe cases, a rapidly progressing lower respiratory tract infection, pneumonia, acute respiratory failure, ARDS, sepsis, and septic shock are often observed. Acute respiratory distress syndrome (ARDS) has been reported to develop in 34% of patients. Hypoxemia (a decrease in SpO2 below 88%) develops in more than 30% of patients. The most severe dyspnea develops on the 6th-8th day after the infection.

**Previous clinical experience** in managing patients with SARS associated with SARS-CoV and MERS coronaviruses defines generally accepted approaches to the treatment of COVID-19 that consist of proactive treatment prior to the development of a complete complex of life-threatening symptoms. Medical care provided to the patients includes monitoring to detect signs of worsening in their clinical condition. Patients infected with SARS-CoV-2 receive supportive pathogenetic and symptomatic treatment. Several drugs are outlined for the treatment of COVID-19: chloroquine, hydroxychloroquine, mefloquine, lopinavir+ritonavir, tocilizumab, azithromycin (in combination with hydroxychloroquine), and recombinant human interferons β-1b and α-2b. Drugs in the clinical trial phase for the treatment of patients with COVID-19 include umifenovir, remdesivir, and favipiravir. However, the currently available information on the results of treatment with all of these drugs does not provide an unambiguous conclusion about their effectiveness or ineffectiveness.

**The antiviral mechanisms of medications** that are considered promising in the treatment of COVID-19 mainly include nonspecific (inhibition or activation) or specific (inhibition) effects on the immune system: 1) antimalarial drugs - nonspecific inhibition of the biosynthetic processes of proteins and RNA/DNA by increasing the lysosomal pH in both epithelial cells and immunocompetent cells that can inhibit the penetration of the virus into the cell and its replication and determines the anti-inflammatory and immunosuppressive effects; 2) antiretroviral drugs - nonspecific inhibition of cellular RNA/DNA biosynthesis by inhibiting reverse transcriptase, proteases, and other enzymes in the cells and tissues of the body that are characterized by active proliferation; 3) interferon drugs - nonspecific activation of cellular and humoral immunity with an increase in cytokine production, suppression of proliferation and acceleration of apoptosis in immunocompetent cells; 4) tocilizumab - a specific inhibition of IL-6 receptors for the rapid and effective suppression of immunity in order to eliminate the life-threatening symptoms of the cytokine release syndrome in patients with a moderate to severe disease state.

**A significant side effect** of antimalarial drugs is cardiotoxicity. A randomized controlled trial showed that the treatment of a disease caused by SARS-CoV-2 with lopinavir+ritonavir monotherapy did not shorten the duration of hospitalization and was not more efficient than regular symptomatic therapy. Before the etiological diagnosis is confirmed, it is recommended to include intranasal forms of interferon-α2b, interferon-inducing agents, and broad-spectrum antiviral drugs, such as umifenovir, in the treatment. However, objective evidence of the efficacy of the above medicinal products in the case of COVID-19 is missing. The use of medicinal products specifically blocking pro-inflammatory cytokines requires cautious consideration of the risk-to-benefit ratio for the patient.

Many countries are currently working on the development of several types of vaccines for **specific prophylaxis of COVID-19**; nevertheless, at present, there are no approved drugs. Nonspecific preventive activities mainly comprised preventing the importation and spread of COVID-19 and are applied to the source of infection (a sick person), the mechanism of pathogen transmission, and the potentially susceptible contingent (protection of individuals who are and/or were in contact with a sick person).

**Pre-exposure prophylaxis** of COVID-19 is recommended as an intranasal administration of recombinant interferon-α2b for healthy individuals. For postexposure prophylaxis in individuals with a single contact with a confirmed case of COVID-19, as well as in individuals in a focus of infection, pharmaceutical prophylaxis with chloroquine, hydroxychloroquine, and mefloquine is recommended, despite their cardiotoxicity. There are currently no proven protocols for effective therapeutic prophylaxis of COVID-19.

**From the pathophysiological point of view**, for the development and application of efficient prevention and treatments of COVID-19, a number of previously published data on the genetic and molecular mechanisms of SARS-CoV-2 interaction with target cells are important. According to these data, a main step of COVID-19 pathogenesis is assumed to be the dose and duration of the viral effect on ventilation-perfusion regulation of the peripheral parts of the bronchopulmonary system rather than virus replication itself with epithelial cell damage and an induced immune system response.

**J.K. Millet & G.R. Whittaker (2018)** proposed a model for the penetration of a cell by SARS-CoV and coronaviruses in general. The model combines various activators and triggers of the viral S-protein as follows: 1) S-protein cleavage by a protease at the binding site of S1 and S2 domains, which provides a preliminary stage of priming; 2) receptor interaction; 3) cleavage of the S2 domain by a protease to release viral fusion peptides; 4) ion changes that contribute to conformational changes and insertion of fusion peptides, caused not only by H+ ions but also by $Ca^{2+}$ ions, which are necessary for fusion peptide insertion into the lipid bilayer of the cell membrane.

**A comparative study of genomes of the viruses** identified 380 amino acid substitutions between SARS-CoV-2 and SARS-CoV (SARS-like coronaviruses), which may have caused a functional and pathogenic divergence of SARS-CoV-2 (Wu A. et al., 2020). A similarity was found between the external part of the SARS-CoV-2 and SARS-CoV S-protein S1 domains, suggesting that these viruses can use membrane ACE2 as a penetration point into the target cell (Lu R. et al., 2020). Furthermore, it was confirmed by cryo-electron microscopy and surface plasmon resonance methods that both viruses use the ACE2 receptor to enter the cell and employ the serine protease TMPRSS2 with ATP for S-protein priming (Hoffmann M. et al., 2020; Wrapp D. et al., 2020). Using molecular simulation methods, it was calculated that ACE2 has two orders of magnitude greater binding to SARS-CoV-2 than to SARS-CoV (Ortega J.T. et al., 2020).

**ACE2 is a membrane enzyme** with an active center located on the surface of type 2 alveolar cells, arterial and venous endothelial cells, smooth muscle cells of the lungs and many other organs (Hamming I. et al., 2004). It mainly acts as a counterweight to the angiotensin-converting enzyme (ACE), which catalyzes the conversion of angiotensin I into the vasoconstrictor hormone angiotensin II. In this regard, it is interesting that in recent clinical observations of patients with severe COVID-19, a high incidence of concomitant diseases such as hypertension and type I or type II diabetes was noted; for the treatment of these diseases, inhibitors of circulating ACE and/or angiotensin I receptor antagonists are used, which increase the expression of ACE2 (Fang L., Karakiulakis G., Roth M., 2020), and this, in turn, can increase the susceptibility of such patients to infection with the SARS-CoV-2 virus.

**Based on the known data** on the physiological role of ACE2 and the extremely high affinity of the SARS-CoV-2 virus, we can assume the following picture of the pathogenesis of this viral infection that defines not only the clinical course of COVID-19 but also the most effective means and methods of active pharmaceutical prophylaxis of this infectious disease. When the SARS-CoV-2 virus contacts the mucous membrane of the tracheobronchial tree, the SARS-CoV-2 viral envelope glycoprotein has a high-affinity interaction with the ACE2 active centers on the apical surface of all available cells, including epithelial cells, alveolar cells, alveolar macrophages, and other cellular elements. Further stages of virus penetration, including immersion of the SARS-CoV-2 + ACE2 complex inside the cell and the beginning of viral RNA replication, are virtually determined and relatively fast processes. Moreover, the amount of the viral load at the time of infection will largely determine the degree of simultaneous functional suppression of ACE2 bronchodilating and vasodilating activity and the relative functional prevalence of ACE constrictor activity in the walls of the bronchi and alveoli.

**The biosynthesis of all membrane proteins** in cells is continuous, but it cannot be replenished earlier than within a few hours after infection with SARS-CoV-2, during which peripheral parts of the lungs will exhibit increased hypoxic processes and disturbances in the ventilation and perfusion properties associated with a reduction in the smooth muscle cells of blood vessels and bronchi. The outcome of subsequent pathogenetic events will largely depend on the duration and reversibility of physiological changes at this stage. Thus, incomplete restoration of ACE2 expression after infection with SARS-CoV-2 with continued synthesis and dissemination of viral particles alongside vaso- and bronchoconstriction during the incubation period will determine a further increase in hypoxic and metabolic changes in the anatomical structures of the blood-air barrier, including the basement membrane and interstitial tissue, thus enhancing the constrictive effects and closing the "vicious circle" of the pathogenesis of destructive inflammatory changes in the lung tissue. Even with a clinically asymptomatic course of the infectious process, these changes, increasing in the peripheral parts of the lung at 7-14 days or earlier, can cause respiratory failure and a decrease in pneumatization of the lung tissue manifested as ground-glass opacity on radiological images, as well as other symptoms characteristic of typical pneumonia with COVID-19.

**The potential efficiency** of vasodilating agents against the virus can be confirmed by the results of clinical trials of camostat mesylate, a serine protease TMPRSS2 inhibitor, which lowers blood pressure by reducing the activity of protease inhibition of epithelial sodium channels (Kitamura K., Tomita K., 2012), efficiently blocks the penetration of the virus into the cell, and is therefore considered a potential antiviral treatment option for COVID-19 (Hoffmann M. et al., 2020). In addition, data on possible inhibition of the penetration of the SARS-CoV virus into cells by the antiarrhythmic medication amiodarone appear to be relevant (Stadler K. et al., 2008).

**For effective pharmaceutical prophylaxis of COVID-19,** among the medicinal products that appear to be promising for application in healthy individuals, combination medicinal products that improve metabolic and redox processes in the cells of the bronchopulmonary system, exhibit antihypertensive and bronchodilating properties and are used in minimally effective doses to exclude possible toxic and side effects should be considered. Examples of these agents are antioxidants (glutathione, ubiquinone, superoxide dismutase), metabolic precursors and regulators of ATP synthesis (inosine, nicotinamide), direct or indirect regulators of ion channels in the membranes of epithelial and smooth muscle cells (potassium and/or magnesium medicinal products, furosemide), β-adrenergic agonists, myotropic antispasmodics, and inducers of the endogenous synthesis of nitric oxide.

**To reduce the clinical manifestations** after the contact of the SARS-CoV-2 virus with the cells of the tracheobronchial tree, as well as to increase the efficacy of local exposure and minimize the systemic side effects of medicinal products in healthy individuals, the aerosol administration of aqueous solutions of medicinal products appears to be the most appropriate method of application. For example, it was previously shown in observational studies on healthy volunteers that inhaled furosemide in low concentrations (up to 40 mg) has a relaxing effect on smooth muscle cells of the airways of the human lungs due to the same mechanism as in the cells of the loop of Henle (Bianco S. et al., 2000; Inokuchi R. et al., 2014; Spicuzza L. et al. 2003; Cavliere F., Masieri S., 2002; Masoumi K. et al., 2014) and may also exhibit immunostimulant properties (Bialasiewicz P. et al., 2004).

### 2. STUDY ASSESSMENT

#### 2.1. Objective

To assess the efficacy and safety of aerosol combination therapy for the pre-exposure prophylaxis of COVID-19 in healthy volunteers at high risk of exposure to SARS-CoV-2 infection.

#### 2.2. Endpoints

3.2.1 Determine the frequency of laboratory confirmed SARS-CoV-2 cases, defined by RT-PCR positivity within 28 days observational period, in high risk population with routine COVID-19 positive contacts.

3.2.2 Evaluation of the safety of aerosol combination therapy as the pre-exposure prophylaxis for COVID-19.

#### 2.3. Design

The study is intended as a low-interventional prospective open-label clinical study of aerosol combination therapy in a cohort of healthy adult volunteers regularly exposed to COVID-19 positive contacts.

#### 2.4. Population

This is an open-label study is intended to obtain preliminary estimates in healthy adults of the efficacy and safety of the combination medication. Based on the observational trials in high risk individuals exposed to routine COVID-19 positive contacts frequency of SARS-CoV infection confirmed by RT-PCR is 11.1% (García I.S. et al., 2020). We expect that the investigated pre-exposure prophylaxis allows to eliminate SARS-CoV-2 positive cases to 3% or less during observational period. With Alpha error rate of 0.05, beta error 0.85, we need to include 96 subjects.

Rare adverse events (AEs) are not demonstrable in a clinical study of this size. With the assumption that all enrolled subjects will likely complete the treatment and safety visits in this relatively short duration study, the following statistical considerations apply. With 100 subjects in the treatment group, the chance of observing at least one AE of probability 5% or more is approximately 95%. Therefore, if no AEs of a given type occur, we can be relatively confident that they will occur in fewer than 5% of people.

#### 2.5. Inclusion Criteria

2.5.1. Healthy volunteers aged between 18 and 80 years;

2.5.2. Informed consent to participate in the study signed by a healthy volunteer;

2.5.3. An ability to cooperate adequately (the volunteer can understand the information provided about the study and is willing to comply with the requirements of the study protocol);

#### 2.6. Exclusion Criteria

Healthy volunteers cannot be included in the trial if they meet at least one of the following criteria.

2.6.1. Criteria related to the disease:

Detection of SARS-CoV-2 virus RNA by polymerase chain reaction (PCR) in biomaterial samples and/or positive enzyme-linked immunosorbent assay ELISA IgM and ELISA IgG to the virus in a healthy volunteer;

2.6.2. Criteria related to the combination medication, previous therapy and concomitant therapy:

Hypersensitivity or individual intolerance to the components of the combination therapy according to the medical history.

2.6.3. Criteria related to the concomitant pathology:

According to the medical history and physical examination:
- any diseases or conditions that, in opinion of the investigator, would make it difficult to interpret the results of treatment or may limit the participation of a healthy volunteer in the study.
According to screening laboratory tests:
- related to the compliance of healthy volunteers in the study;
- any diseases or conditions that, in opinion of the investigator, would make it difficult to interpret the results of treatment or may limit the participation of healthy volunteers in the study;
Healthy volunteers who, in the opinion of the investigator, are obviously or likely unable to understand and evaluate the information regarding this study within the process of signing the informed consent form, in particular regarding the expected risks and possible discomfort.
The inability or unwillingness of a healthy volunteer to follow the rules for carrying out the study and participating in a clinical trial.

#### 2.7. Early Discontinuation Criteria

2.7.1. A healthy volunteer will be excluded from the study if SARS-CoV-2 virus RNA is detected in biomaterial samples by a polymerase chain reaction (PCR) for COVID-19 or if enzyme-linked immunosorbent assay (ELISA) IgM and ELISA IgG results are positive.

2.7.2. A healthy volunteer may withdraw their consent to participate in the study at any time and without giving reasons. The investigator may also decide on early discontinuation of the participation of a healthy volunteer in the study at any time if the patient's condition requires it.

2.7.3. Main reasons for early discontinuation of participation of a healthy volunteer in the study:

Individual intolerance to the combination medication or any of its components;
Failure to meet the inclusion and exclusion criteria;
Refusal of a healthy volunteer to participate in the study or the withdrawal of informed consent by a healthy volunteer;
Development of adverse events that prevent further prophylactic therapy with the medication;
The emergence of a serious adverse event in a healthy volunteer;
The emergence of exclusion criteria in the course of the study;
Any condition of a healthy volunteer that requires, in the opinion of the investigator, the withdrawal of the patient from the study;
Discontinuation of the trial by the research center, investigator, or regulatory authority.

#### 2.8. Duration

The trial comprises the following stages:
The screening period lasts up to 2 days.
The treatment period lasts 14 days.
The follow-up period lasts 14 days.
The duration of the study for healthy volunteers will not exceed 30 days.
All visits can only take place in a specialized hospital.

### DESCRIPTION OF THE COMBINATION MEDICATION

**INN:** POTASSIUM CHLORIDE

**Brand names of the pharmaceuticals:** various

**Therapeutic classification:** therapeutic drug for electrolyte balance

**ATC code:** B05XA01 Potassium Chloride

**Composition and pharmaceutical form:**
Concentrate for solution for infusion, a clear, colorless or slightly yellowish liquid in ampules of 10 ml, 10 ampules in a package.
One milliliter of the solution contains the following:
Active substance: potassium chloride – 40 mg
Excipients: Dextrose monohydrate - 334 mg, hydrochloric acid solution 1 M - up to pH 3.0-4.0, DI water - up to 1 ml.

**Pharmacokinetics**
After intake, the drug is easily and passively absorbed (70%) in virtually any amount because its concentration (both from food and released by pharmaceuticals) is higher in the lumen of the small intestine than in the blood. In the ileum and colon, potassium is excreted into the intestinal lumen following the principle of conjugated exchange with sodium and excreted with bile (10%). $T_{1/2}$ in the absorption phase is 1.31 hours.

**Pharmacological effect**
Potassium deficiency replenishing agent. Helps in maintaining the necessary intra- and extracellular levels of potassium. Potassium is the main intracellular ion and has an important role in the regulation of various body functions. It participates in maintaining intracellular osmotic pressure, nerve impulse conduction and transmission to innervated organs, contraction of skeletal muscles, and a number of biochemical processes. Reducing the excitability and conductivity of the myocardium at high doses suppresses automatism.

**Indications for the use of drug**
Hypokalemia of various origins, including vomiting, diarrhea, hyperaldosteronism, polyuria in chronic renal failure, and the use of certain drugs; arrhythmias, including glycoside intoxication; hypokalemic form of paroxysmal myoplegia.

**Contraindications:** impaired renal function, complete heart block.

**Side effects:** nausea, vomiting, diarrhea.

**Registered administration and dose:** iv infusion at a rate of 20-30 drops per minute.

**There are no data on the inhaled administration of the drug.**

**Manufacturers - owners of the registration license:**
GROTEX OOO (Russia), SINTEZ OAO (Russia), BINNOFARM AO (Russia) and others.

VIDAL 22.04.2020 https://www.vidal.ru/drugs/potassium_chloride__44667

**INN:** INOSINE GLYCYL-CYSTEINYL- GLUTAMATE DISODIUM

**Brand names of pharmaceuticals:** Molixan®

**Therapeutic classification:** metabolic agent

**ATC code:** V03AX Other therapeutic products

**Composition and pharmaceutical form:** solution for intravenous and intramuscular administration; 1 ml of a 3% solution for injections contains inosine glycyl-cysteinyl-disodium glutamate - 30 mg; in ampules of 1 and 2 ml with a break ring or dot; 5 or 10 ampules in a blister pack, 1 blister in a cardboard box.

One milliliter of the solution contains the following:
Active substance: inosine glycyl-cysteinyl-disodium glutamate 30 mg.
Excipients: sodium acetate trihydrate 13.6 mg, diluted acetic acid up to pH 6.0, and water for injections up to 1 ml.

**Pharmacological effect:** antiviral, immunomodulatory, hepatoprotective.

**Mechanism of action:** regulates the thiol-disulfide exchange of epithelial and immunocompetent cells, the production of cytokines by macrophages, including interleukin-2, α- and γ-interferons, inhibits the cytolysis of functionally active cells and induces apoptosis of virus-infected cells.

**Contraindications:** individual hypersensitivity, pregnancy, breastfeeding.

**Side effects:** uncommon - increased body temperature (up to 37.1–37.5°C), slight soreness at the injection site.

**Registered administration and dose:** iv, i/m for lasting and severe forms of acute viral hepatitis B: 10–30 mg/day daily, single-dose, duration of the course: 24 days (in combination with symptomatic and detoxification therapy). In the case of chronic viral hepatitis B: 10–30 mg/day, daily, single-dose, for 3 months or more (combination with antiviral chemotherapy is possible). In the case of chronic viral hepatitis C: 30–60 mg/day in 1-2 doses, treatment duration 3-6 months (in combination with antiviral chemotherapy).

**There are no data on the inhaled administration of the drug.**

**Owner of the registration license:** ZAO PharmaVAM (Russia)

**Manufacturers:** State Research Institute of Highly Pure Biopreparations of the Federal Medical-Biomedical Agency, Russian Cardiology Research and Production Complex of the Ministry of Health of Russian Federation

VIDAL 22.04.2020 https://www.vidal.ru/drugs/molixan__30093

### 3. DOSAGE AND ADMINISTRATION OF A COMBINATION MEDICATION

The combination medication is administered daily in the daytime 4 (four) times a day with an interval of at least 4 hours.

The combination of the medication is prepared per daily inhalation dose by mixing the following medication in the indicated amount *ex tempore*: 3% solution of inosine glycyl-cysteinyl-disodium glutamate 4.0 ml and 4% potassium chloride solution 1.0 ml.

The combination medication should be used within 24 hours after preparation.

One milliliter of a solution of a combination medication contains active substances:
Potassium chloride 8.0 mg
Inosine glycyl-cysteinyl-glutamate disodium 24.0 mg

A single inhalation dose of the combination medication is 1.25 ml.

The combination medication for inhalation is administered using a nebulizer equipped with a mouthpiece. The nebulizer is connected to the compressor to create the required air flow (5-8 L/min), and the nebulizer must be filled with at least 4 ml of the medicinal product.

The combination medication should be inhaled through the mouthpiece of the nebulizer in a calm and regular manner.

The nebulizer should be regularly cleaned following the manufacturer's instructions. Clean the cup of the nebulizer after each use. Wash the cup of the nebulizer and mouthpiece or mask with warm water and mild detergent (follow the manufacturer's instructions). The nebulizer should be thoroughly rinsed and dried by connecting the cup to the compressor or air inlet valve.

**NB! Do not use ultrasonic nebulizers for the administration of the combination medication**

### 4. CLINICAL AND LABORATORY ASSESSMENT

Clinical and laboratory examination of volunteers within 48 hours before the beginning of combination medication usage.

#### 4.1. **Complaints Assessment.**

#### 4.2. **Clinical Examination:**

Physical examination (assessment of the visible mucous membranes of the upper respiratory tract, auscultation and percussion of the lungs, palpation of the lymph nodes, abdominal examination and determination of the size of the spleen and liver, thermometry, BP, HR, RR.

#### 4.3. **Laboratory Tests:**

Detection of SARS-CoV-2 coronavirus RNA by RT-PCR in accordance with the "Interim Guidelines of the Ministry of Health of the Russian Federation on the Prevention, Diagnosis, and Treatment of the Novel Coronavirus Infection COVID-19, Version 5 (08.04.2020)".

### 5. BENEFIT-RISK ASSESSMENT

#### 5.1. Efficiency:

The efficacy of the therapy will be assessed based on the analysis of the primary outcome - a development rate among healthy volunteers of the infectious process caused by the SARS-CoV-2 virus during the study, as confirmed by the detection of viral RNA in biological material using PCR according to the Temporary Guidelines of the Ministry of Health of the Russian Federation on the prevention, diagnosis, and treatment of the novel coronavirus infection COVID-19.

#### 5.2. Safety:

The rate and intensity of the clinical manifestations of adverse events (AE).

The rate and intensity of deviations from the reference values of laboratory parameters and instrumental data

##### 5.2.1. Methods and time schedule for assessment, registration, and analysis of safety parameters

The development rate and intensity of AE will be assessed in the physical examination at each visit, as well as during and after each administration of the medicinal product.

The conclusion about the safety of the administered medication will be made after a statistical evaluation of all AEs, including serious AEs, which have revealed at least a possible relation with the administration of the pharmaceuticals used.

##### 5.2.2. Adverse events registration and reporting

<u>Adverse events (AE)</u>

The protocol requires the registration of all AEs that occur in the patient during and after administration of the medicinal product and until the end of the patient's participation in the trial.

An adverse event is defined as “any untoward medical occurrence in a patient in a clinical investigation who received a medicinal product; the event does not necessarily have to have a causal relationship with this treatment” (National Standard of the Russian Federation GOST R52379-2005 “Good Clinical Practice”).

An AE can be any symptom (including a clinically significant deviation of the laboratory measurement from the reference values), complaint, or disease, the time of occurrence of which does not exclude a causal relationship with the use of the investigational medicinal product, regardless of the existence of such a relationship.

AEs are classified by severity (nonserious or serious AEs), causal relationship with the investigational medicinal product (no/yes; if yes: probable, possible, unlikely, conditional, unclassifiable), and intensity (mild, moderate, severe).

<u>Serious adverse events (SAEs)</u>

In the framework of this protocol and in accordance with legal requirements (National Standard of the Russian Federation GOST R52379-2005 “Good Clinical Practice” dated September 25, 2005), a SAE is defined as any adverse medical event that, regardless of the dose of the medicinal product, results in death, is immediately life-threatening, requires or prolongs patient hospitalization, results in a persistent or significant disability or incapacity, or is a congenital anomaly or birth defect.

Intensity of AE

The intensity of the AE should be judged based on the following classification:

- Level 1 - Mild: an adverse event that is easily tolerated by the patient, causing minimal inconvenience and not interfering with the patient's daily activities.
- Level 2 - Moderate: an adverse event that causes discomfort and interferes but does not impede the patient's daily activities.
- Level 3 - Severe: an adverse event that impedes normal daily activities.

Causal relationship between AEs and the investigational medicinal product:

The investigator will evaluate the relationship between the development of an adverse event and the investigational medicinal product as follows:
No - it is clearly and unconditionally related only with extraneous reasons and does not meet the criteria for an unlikely, possible, or probable relationship.
Yes - there is a reasonable causal relationship with the administered investigational product

Criteria:

- could have been caused by a clinical state or external factors or other prescribed treatments;
- a clear temporal relationship between discontinuation of the investigational medicinal product or dose reduction and improvement;
- recurrent upon repeated administration;
- corresponds to the known response to the investigational medicinal product.

The relationship between AEs and the investigational medicinal product will be assessed by a WHO scale, considering the design of the study.

Certain. Clinical manifestations of AEs and laboratory test abnormalities appear during the medicinal product intake and cannot be explained by diseases or other factors.

Probable. Clinical manifestations of AEs and laboratory test abnormalities have a reasonable time relationship to the medicinal product intake and are unlikely to be attributed to concomitant diseases or other factors.

Possible. Clinical manifestations of AEs and laboratory test abnormalities have a reasonable time relationship to the medicinal product intake; however, they could also be explained by concomitant diseases or the use of other pharmaceuticals and the influence of chemical compounds.

Unlikely. Clinical manifestations of AEs and laboratory test abnormalities have an improbable (but not impossible) time relationship to medicinal product intake; other factors (pharmaceuticals, diseases, chemical compounds) provide plausible explanations for the events.

Conditional. Clinical manifestations of AEs and laboratory test abnormalities related to the AEs are difficult to evaluate. More data for proper assessment are needed, or these additional data are under examination.

Unclassifiable. Reports suggesting an AE cannot be judged because the information is insufficient or contradictory.

Terms used with respect to the investigational medicinal product:

An adverse event (“side effect”) means any untoward medical occurrence in the subject of the study that is associated with the use of the registered medicinal product in the doses recommended in the instructions for medical use of the product.

Unanticipated adverse event - an adverse event, the nature or severity of which is not consistent with known product information, for example, with instructions for medical use in the case of a registered medicinal product.

Severe unanticipated adverse event (SUAE) - an unanticipated adverse event that is characterized by features of SAE.

Registering adverse events

Responsibility for registering AEs in a clinical trial lies with the research doctor. At each visit, the researcher registers all objectively observed or subjectively described AEs in primary documents and individual patient records, filling out the relevant pages of individual patient records and informing the classification of AEs (severity, intensity, relationship with the administration of the medicinal product).

Any available information related to the registered AE, for example, the results of diagnostic tests (laboratory tests, ECG, etc.) should also be attached to the individual patient's record.

Reporting adverse events/serious adverse events

In the case of an AE, the investigator should describe it in as much detail as possible in the primary documentation of the patient, register the AE in the individual patient's record, and fill in the AE Registration Form (Appendix to the individual patient's record).

The following shall be registered in the documents: the clinical manifestation of the AE, the time of occurrence and resolution of the AE, the severity/intensity of the AE, an unanticipated or previously described/known AE, possible relationship with the investigational medicinal product/benchmarking medicinal product administration, relationship with the administration method of the medicinal product, measures taken in relation to the patient and use of the pharmaceutical, drug therapy for the AE, and outcome/resolution of the AE.

Upon receipt of additional information on the development/resolution of the AE, the investigator should register it in the primary documentation and in the previously launched AE Registration Form.

In the case of emerging new symptoms that are not associated with a previously emerged AE, the investigator should launch a new AE form.

In case of the development of an SAE, it should also be registered in the primary documentation and the patient’s individual record; the SAE Registration Form shall be filled out (Appendix to the Individual patient's record).

All AE/SAE Registration Form fields should be filled in. In the absence of information, the N/D field is checked (no data, that is, unknown); alternatively, a dash is drawn in the field (for example, in the "date" field), or a record is made that a given procedure was not performed.

### 6. STATISTICAL ANALYSIS

The methods for statistical analysis of the obtained data are determined by the nature, type, and distribution of the data. The applicability of a number of statistical methods will be evaluated upon the completion of the data collection due to the unknown distribution, sample uniformity, etc. In the course of the analysis, the list of methods used can be expanded if necessary for high-quality data processing.

Statistical processing of data obtained in the study will be carried out using statistical programs or other special software that would ensure high quality of the results. Interval (quantitative) data will be described using the arithmetic mean, standard deviation, median, lower (25%) and upper (75%) quartiles, minimum and maximum values. Categorical (qualitative) data will be described using absolute and relative frequencies. To compare normally distributed quantitative data, standard parametric criteria shall be employed: Student's t-test for dependent/independent samples and analysis of variance (ANOVA) for repeated measurements. For the comparison of quantitative data that are not normally distributed, standard nonparametric tests shall be employed. The frequencies of the variables will be compared between the groups using Pearson's $\chi^2$ test or Fisher's exact test.

AEs will be analyzed based on the frequency of adverse events/serious adverse events. Adverse events registered within the study will be presented by incidence rate (the number of patients with AEs and the number of such AEs in the group). AEs will also be presented by their intensity and relationship with the administration of the combined medicinal product.

Regardless of the reason for the termination of the study, the data of all enrolled healthy volunteers will be taken into account when analyzing the efficiency and safety of the investigational therapy.

### 7. REFERENCES

Bialasiewicz P., Wlodarczyk A., Dudkiewicz B., Nowak D. (2004) Inhibitory effect of furosemide on activation of human peripheral blood polymorphonuclear leukocytes stimulated with n-formyl-methionyl-leucyl-phenylalanine. **International Immunopharmacology**. 4(6):819-931.

Bianco S., Robuschi M., Vaghi A., Fumagalli A., Sestini P. (2000) Inhaled transmembrane ion transport modulators and nonsteroidal anti-inflammatory drugs in asthma. **Thorax.** 55 (Suppl 2):S48-S50.

Cavaliere F., Masieri S. (2002) Furosemide protective effect against airway obstruction. **Current Drug Targets.** 3(3):197-201.

Fang L., Karakiulakis G., Roth M. (2020) Are patients with hypertension and diabetes mellitus at increased risk for COVID-19 infection? **Lancet respiratory medicine**. 8(4):e21.

García I.S., Aramayona López M.J.M., Vicente A.S., Abascal P.L. (2020) SARS-CoV-2 infection among healthcare workers in a hospital in Madrid, Spain. **Journal of Hospital Infection** (in press).

Hamming I., Timens W., Bulthuis M.L., Lely A.T., Navis G., van Goor H. (2004) Tissue distribution of ACE2 protein, the functional receptor for SARS coronavirus. A first step in understanding SARS pathogenesis. **Journal of Pathology.** 203(2):631-637.

Hoffmann M., Kleine-Weber H., Schroeder S., Kruger N., Herrler T. et al.(2020) SARS-CoV-2 Cell Entry Depends on ACE2 and TMPRSS2 and Is Blocked by a Clinically Proven Protease Inhibitor. **Cell.** 181(2):271-280.e8.

Inokuchi R., Aoki A., Aoki Y., Yahagi N. (2014) Effectiveness of inhaled furosemide for acute asthma exacerbation: a meta-analysis. **Critical Care.** 18(6):621.

Kitamura K., Tomita K. (2012) Proteolytic activation of the epithelial sodium channel and therapeutic application of a serine protease inhibitor for the treatment of salt-sensitive hypertension. **Clinical and Experimental Nephrology.** 16(1):44-48.

Lu R., Zhao X., Li J., Niu P., Yang B. et al.(2020) Genomic characterization and epidemiology of 2019 novel coronavirus: implications for virus origins and receptor binding. **Lancet.**395(10224):565-574.

Masoumi K., Forouzan A., Haddadzadeh Shoushtari M., Porozan S., Feli M., Fallah Bagher Sheidaee M., Asgari Darian A. (2014) The efficacy of nebulized furosemide and salbutamol compared with salbutamol alone in reactive airway disease: a double-blind randomized, clinical trial. **Emergency Medicine International.** 2014:638102.

Millet J.K., Whittaker G.R. (2018) Physiological and molecular triggers for SARS-CoV membrane fusion and entry into host cells. **Virology.** 517:3-8.

Ministry of Health of the Russian Federation. The prevention, diagnosis, and treatment of the novel coronavirus infection (COVID-19). Temporary guidelines, Version 5 (08.04.2020), 115p. https://www.rosminzdrav.ru/ministry/med_covid19.

Ortega J.T., Serrano M.L., Pujol F.H., Rangel H.R. (2020) Role of changes in SARS-CoV-2 spike protein in the interaction with the human ACE2 receptor: An in silico analysis. **EXCLI Journal.** 19:410-417.

Spicuzza L., Ciancio N., Pellegrino R., Bellofiore S., Polosa R., Ricciardolo F.L., Brusasco V., Di Maria G.U. (2003) The effect of inhaled furosemide and acetazolamide on bronchoconstriction induced by deep inspiration in asthma. **Monaldi Archives for Chest Disease.** 59(2):150-154.

Stadler K., Ha H.R., Ciminale V., Spirli C., Saletti G., Schiavon M., Bruttomesso D., Bigler L., Follath F., Pettenazzo A., Baritussio A. (2008)Amiodarone alters late endosomes and inhibits SARS coronavirus infection at a postendosomal level. **American Journal of Respiratory Cell and Molecular Biology.** 39(2):142-149.

Wrapp D., Wang N., Corbett K.S., Goldsmith J.A., Hsieh C.L., Abiona O., Graham B.S., McLellan J.S. (2020) Cryo-EM structure of the 2019-nCoV spike in the prefusion conformation. **Science**. 367(6483):1260-1263.

Wu A., Peng Y., Huang B., Ding X., Wang X., Niu P., Meng J., Zhu Z., Zhang Z., Wang J., Sheng J., Quan L., Xia Z., Tan W., Cheng G., Jiang T. (2020) Genome Composition and Divergence of the Novel Coronavirus (2019-nCoV) Originating in China. **Cell Host & Microbe.** 27(3):325-328.

### 8. APPENDICES

Appendix 1

#### Study Plan

| Procedures | Screening | Treatment | | | Follow-up | |
|---|---|---|---|---|---|---|
| **Visit** | **0** | **1** | **2** | **3** | **3** | **4** |
| **Days** | -2 - 0 | 1 | 7 | 14 | 21±1 | 28±1 |
| Informed Consent | X | - | - | - | - | - |
| Insurance | X | | | | | |
| PCR for SARS-CoV-2 RNA | X | X | X | X | X | X |
| ELISA IgM and IgG to SARS-CoV-2 | X | - | - | | - | X |
| Demographics, Complaints and Medical History, Anthropometric Data | X | X | - | - | - | - |
| Inclusion and Exclusion Criteria Assessment | X | X | X | X | X | X |
| Physical Examination | X | X | X | X | X | X |
| Vital Signs Assessment (BP, HR, RR, TEMP) | - | X | X | X | X | X |
| Exclusion Criteria Assessment | X | - | - | - | - | - |
| Previous Therapy Assessment | X | X | X | X | X | X |
| Concomitant Therapy Assessment | X | X | X | X | X | X |

Appendix 2.

### Informed Consent Form

I, ____________________________________________, residing at: ____________________________, give my consent to participate in a low-interventional, open-label prospective study in accordance with the protocol "**MIBVD-19 Low-Interventional Study of the Aerosol Use of a Combination Medication as Pre-Exposure Prophylaxis for Novel Coronavirus Infection COVID-19**" (referred to as - Study), conducted at ______________________________________________________________________________.

I have received thorough clarifications from the principal investigator, who has discussed with me the question of my participation in the study, regarding the nature, purpose, and duration of this study.

I confirm that I have fully read the provided information. I have been given comprehensive and clear information for participation in the study, and the responsibilities of participants in the study have been explained to me. I have had the chance to ask questions, and I am satisfied with the answers.

I understand that participation in this study is voluntary. I can withdraw my consent at any time and without giving reasons, and the decision to leave the study or refusal to participate in the study will not affect the amount and quality of medical care I receive.

I understand that I will be immediately informed of new drug safety data that may affect my consent to continue participation in the study.

I understand that authorized representatives of regulating organizations and the ethics committee can access some sections of my medical records related to my participation in this study. By signing this form, I grant them the right to access my medical records, provided that they maintain professional confidentiality.

I confirm that at the time of signing this consent, I am not a pregnant or breastfeeding woman, in order to completely exclude the risk of harm occurring during pregnancy or breastfeeding to me, the fetus, or the baby.

I understand that this study will collect information that will be treated as confidential.

I will not try to limit the possible use of the research results.

I agree to participate in this study and collaborate with the principal investigator ______________________, as well as with authorized members of their group. I undertake to immediately inform the principal investigator of any noticed deviations from the norm.

I agree that the principal investigator may contact my relatives or acquaintances, the attending physician, or other doctors responsible for my treatment to obtain information about my health state, if this is important for this study.

I have been informed that the principal investigator will provide telephone numbers for further contact, and if necessary, at any reasonable time I can inform the investigator of a change in my health status or ask a question.

I have received a signed copy of this informed consent form.

By signing this form, I confirm that I am willing to participate in the study.

| ____________________ | ____________________ | ____________________ |
|---|---|---|
| *Volunteer name, surname* | *Date* | *Signature* |
| ____________________ | ____________________ | ____________________ |
| *Principal investigator* | *Date* | *Signature* |

Appendix 3

### Instruction for Use of Combination Medication

#### 1. Preparation of Combination Medication

A combination of the medication is prepared per daily inhalation dose by mixing the following medication in the indicated amount *ex tempore*:

3% solution of inosine glycyl-cysteinyl-disodium glutamate - 4.0 ml;
4% potassium chloride solution - 1.0 ml.

One milliliter of a solution of a combination medication contains the following active substances:

Inosine glycyl-cysteinyl-disodium glutamate - 24.0 mg
Potassium chloride - 8.0 mg

#### 2. Dosage and Administration of the Combination Medication

The combination medication is administered daily in the daytime 4 times a day with an interval of at least 4 hours.
The combination medication should be used within 24 hours after preparation.
A single inhalation dose of the combination medication is 1.25 ml.
A single dose of medication of the combination is as follows:

Inosine glycyl-cysteinyl-disodium glutamate – 30.0 mg
Potassium chloride – 10.0 mg

A daily dose of medication of the combination is as follows:

Inosine glycyl-cysteinyl-disodium glutamate – 120.0 mg
Potassium chloride – 40.0 mg

The combination medication is administered by inhalation using a nebulizer equipped with a mouthpiece. The nebulizer is connected to the compressor to create the required air flow (5-8 l/min). The combination medication should be inhaled through the mouthpiece of the nebulizer in a calm and regular manner.
The recommended duration of combined medicinal product use is 14 days.

#### 3. Other Recommendations

The nebulizer will be regularly cleaned following the manufacturer's instructions. The cup of the nebulizer will be cleaned after each use. The cup of the nebulizer and mouthpiece or mask will be washed with warm water and mild detergent (following the manufacturer's instructions). The nebulizer will be thoroughly rinsed and dried by connecting the cup to the compressor or air inlet valve.

#### 4. Special indications

Do not use ultrasonic nebulizers for the administration of the combination medication.

Appendix 4

### Individual Record Form

Protocol: MIBVD-19 Low-Intervention Study of the Aerosol Use of Combination Medication as Pre-Exposure Prophylaxis for the Novel Coronavirus Infection COVID-19.

**Identification:**

Registration Number _ _ _

Surname ________________________________

Name ________________________________

Patronymic ____________________________

Sex **MALE/FEMALE**

Year of birth _ _. _ _. _ _

Full years _ _

**Investigator:**____________________________________ ____________________

*Name Surname* *Signature*

**Date:** «____» ______________ 2020

### **Visit 0** (Screening)
### Study day -2-0

Date _ _. _ _. _ _ _ _

Height _ _ _ (cm)

Weight_ _ (kg)

**Bad habits:**

Smoking: **YES/NO**
If yes, specify how many cigarettes per day ________________________________________

Allergies: **YES/NO**
If yes, specify, to what and how it is manifested ________________________________________

Use of alcohol: **YES/NO**
If yes, specify what kind of alcohol and the weekly amount (ml) ________________________________

**Medical history:**

Cardiovascular diseases: **YES/NO**
If yes, specify ________________________________________________________________

Treatment for cardiovascular diseases: **YES/NO**
If yes, specify which medications and in what dose are taken daily ________________________

Pulmonary diseases: **YES/NO**
If yes, specify ________________________________________________________________

Treatment for pulmonary diseases: **YES/NO**
If yes, specify which medications and in what dose are they taken daily________________________

Gastrointestinal diseases: **YES/NO**
If yes, specify ________________________________________________________________

Treatment for gastrointestinal diseases: **YES/NO**
If yes, specify which medications and in what dose are they taken daily________________________

Musculoskeletal system diseases: **YES/NO**
If yes, specify ________________________________________________________________

Treatment for musculoskeletal system diseases: **YES/NO**
If yes, specify which medications and in what dose are they taken daily ________________________

Diabetes: **YES/NO**

Treatment for diabetes: **YES/NO**
If yes, specify which medications and in what dose are they taken daily________________________

Acute respiratory viral infections or influenza in the last 3 months: **YES/NO**

**Complaints during an examination:** **YES/NO**

If yes, specify, what is present:

Weakness **YES/NO**

Muscular pain **YES/NO**

Headaches **YES/NO**

Dyspnea **YES/NO**

Cough **YES/NO**

Nasal congestion **YES/NO**

Increased temperature **YES/NO**

Sore throat **YES/NO**

Other **YES/NO**

If yes, describe in detail ____________________________________________________________

**Physical examination:**

**Body temperature __ __ °C**

**Swollen lymph nodes:** **YES/NO**

If yes, underline which nodes are affected: occipital, posterior cervical, parotid, anterior cervical, submandibular, sublingual, supraclavicular and subclavian, axillary, epitrochlear, inguinal, popliteal.

**Pharyngeal examination:** hyperemia **YES/NO**

**BP sitting _ _ _/_ _ _**(mm Hg)

**HR _ _ _**per min

**RR _ _** per min

**Wheezing in the lungs (dry/wet):** **YES/NO**
If yes, describe the nature and location
___________________________________________________________

**Abdominal palpation:** soft, painless **YES/NO**

**Liver:** aligned with the costal arch: **YES/NO**
If no, describe in detail ______________________________________________________________________

**Spleen:** not enlarged **YES/NO**
If not, describe in detail ___________________________________________

**Specific laboratory diagnostics:**

**Tested with PCR for SARS-CoV-2 RNA**: **YES/NO**
If yes, indicate the results ___________________________________________

**Current therapy:** **YES/NO**
If yes, describe in detail the pharmaceuticals and daily doses___________________________

**Assessment of adverse events:** **YES/NO**
If yes, describe in detail the nature of the adverse event___________________________

**Assessment of inclusion/exclusion criteria**

**Inclusion criteria**

Healthy volunteers aged between 18 and 80 years: **YES/NO**

Informed consent form signed by a healthy volunteer: **YES/NO**

Ability of a volunteer to cooperate adequately (the volunteer can understand the information provided about the study and is willing to comply with the requirements of the study protocol): **YES/NO**

**Exclusion criteria**

Healthy volunteers cannot be included in the trial if they meet at least one of the following criteria.

<u>Criteria related to the underlying disease</u>:

Detection of SARS-CoV-2 virus RNA by polymerase chain reaction (PCR) for COVID-19 in biomaterial samples and positive reactivity of enzyme-linked immunosorbent assay (ELISA) IgM and ELISA IgG to the virus in a healthy volunteer: **YES/NO**

Criteria related to the combination medication, previous therapy and concomitant therapy: **YES/NO**

Hypersensitivity or individual intolerance to the components of the combination medication products according to the medical history: **YES/NO**

<u>Criteria related to the concomitant pathology</u>:

According to the medical history and physical examination: **YES/NO**

Any diseases or conditions that, in opinion of the investigator, make it difficult to interpret the results of treatment or may limit the participation of a healthy volunteer in the study: **YES/NO**

<u>According to screening laboratory tests:</u>

Related to the compliance of healthy volunteers in the study: **YES/NO**

Any other diseases or conditions that, in the opinion of the investigator, would make it difficult to interpret the results of treatment or may limit the participation of a healthy volunteer in the study: **YES/NO**

Healthy volunteers who, in the opinion of the investigator, are obviously or likely unable to understand and evaluate the information regarding this study within the process of signing the informed consent form, in particular, regarding the expected risks and possible discomfort: **YES/NO**

The inability or unwillingness of a healthy volunteer to follow the rules for carrying out the study and participating in a clinical trial: **YES/NO**

**Based on the assessment of the inclusion/exclusion criteria, the healthy volunteer may be admitted to the study: YES/NO**

If the volunteer continues the trial, the date of the next visit is **_ _. _ _. _ _ _ _**

**Investigator:**_____________________________________________ ________________________

*Name Surname* *Signature*

**Date:** «____» ________________ 2020

### **Visit 1** (Treatment period)
### Study day 1

**Date of the visit** _ _. _ _. _ _ _ _

**Physical examination:**

**Body temperature __ __ °C**

**Swollen lymph nodes:** **YES/NO**

If yes, underline which nodes are affected: occipital, posterior cervical, parotid, anterior cervical, submandibular, sublingual, supraclavicular and subclavian, axillary, epitrochlear, inguinal, popliteal.

**Pharyngeal examination:** hyperemia **YES/NO**

**BP sitting _ _ _/_ _ _**(mm Hg)

**HR _ _ _**per min

**RR _ _** per min

**Wheezing in the lungs (dry/wet):** **YES/NO**
If yes, describe nature and location ______________________________________________

**Abdominal palpation:** soft, painless **YES/NO**

**Liver:** aligned with the costal arch: **YES/NO**
If no, describe in detail ______________________________________________

**Spleen:** not enlarged **YES/NO**
If not, describe in detail ______________________________________________

**Tested with PCR for SARS-CoV-2 RNA**: **YES/NO**
If yes, indicate the results ______________________________________________

**Current therapy:** **YES/NO**
If yes, describe in detail the pharmaceuticals and daily doses______________________________

**Assessment of adverse events:** **YES/NO**
If yes, describe in detail the nature of the adverse event______________________________

**Assessment of inclusion/exclusion criteria**

**Inclusion criteria**

Healthy volunteers aged between 18 and 80 years: **YES/NO**

Informed consent form signed by the healthy volunteer: **YES/NO**

Ability of a volunteer to cooperate adequately (the volunteer can understand the information provided regarding the study and is willing to comply with the requirements of the study protocol): **YES/NO**

**Exclusion criteria**

Healthy volunteers cannot be included in the trial if they meet at least one of the following criteria.

Criteria related to the underlying disease:

Detection of SARS-CoV-2 virus RNA by polymerase chain reaction (PCR) for COVID-19 in biomaterial samples and positive reactivity of enzyme-linked immunosorbent assay (ELISA) IgM and ELISA IgG to the virus in a healthy volunteer: **YES/NO**

Criteria related to the combination medication products, previous therapy and concomitant therapy: **YES/NO**

Hypersensitivity or individual intolerance to the components of the combination medication products according to the medical history: **YES/NO**

Criteria related to the concomitant pathology:

According to the medical history and physical examination: **YES/NO**

Any diseases or conditions that, in opinion of the investigator, would make it difficult to interpret the results of treatment or may limit the participation of a healthy volunteer in the study: **YES/NO**

According to screening laboratory tests:

Related to the compliance of healthy volunteers in the study: **YES/NO**

Any other diseases or conditions that, in the opinion of the investigator, would make it difficult to interpret the results of treatment or may limit the participation of a healthy volunteer in the study: **YES/NO**

Healthy volunteers who, in the opinion of the investigator, are obviously or likely unable to understand and evaluate the information regarding this study within the process of signing the informed consent form, in particular regarding the expected risks and possible discomfort: **YES/NO**

The inability or unwillingness of a healthy volunteer to follow the rules for carrying out the study and participating in a clinical trial: **YES/NO**

**Based on the assessment of the inclusion/exclusion criteria, the healthy volunteer may be admitted to the study: YES/NO**

**Assessment of exclusion criteria**

A healthy volunteer will be excluded from the study if SARS-CoV-2 virus RNA is detected in biomaterial samples by a polymerase chain reaction (PCR) for COVID-19 or if there is positive reactivity to enzyme-linked immunosorbent assay (ELISA) IgM and ELISA IgG: **YES/NO**

A healthy volunteer may withdraw their consent to participate in the study at any time and without giving reasons. The investigator may also decide on early discontinuation of the participation of a healthy volunteer in the study at any time if the patient's condition requires it: **YES/NO**

Main reasons for early discontinuation of participation of a healthy volunteer in the study: **YES/NO**

Individual intolerance to the combination medication products or any of its components: **YES/NO**

Failure to meet the inclusion/exclusion criteria: **YES/NO**

Refusal of a healthy volunteer from participation in the study/withdrawal of informed consent by a healthy volunteer: **YES/NO**

Development of AEs that prevent further prophylactic therapy with the combination medication products: **YES/NO**

Emergence of a serious adverse event in a healthy volunteer: **YES/NO**

The emergence of exclusion criteria in the course of the study: **YES/NO**

Any condition of a healthy volunteer that requires, in the opinion of the investigator, the withdrawal of the patient from the study: **YES/NO**

Discontinuation of the trial by the research center, investigator, or regulatory authority: **YES/NO**

**Patient meets all the criteria of the study: YES/NO**
If not, specify the reason in detail ____________________________________________
________________________________________________________________________________

If the volunteer continues the trial, the date of the next visit is **_ _. _ _. _ _ _ _**

**Investigator:**_________________________________________ ______________________
*Name Surname* *Signature*

**Date:** «____» _______________ 2020

### **Visit 2** (Treatment period)
### Study day 7

**Date of the visit** _ _. _ _. _ _ _ _

**Physical examination:**

**Body temperature __ __ °C**

**Swollen lymph nodes:** **YES/NO**

If yes, underline which nodes are affected: occipital, posterior cervical, parotid, anterior cervical, submandibular, sublingual, supraclavicular and subclavian, axillary, epitrochlear, inguinal, popliteal.

**Pharyngeal examination:** hyperemia **YES/NO**

**BP sitting _ _ _/_ _ _**(mm Hg)

**HR _ _ _**per min

**RR _ _** per min

**Wheezing in the lungs (dry/wet):** **YES/NO**
If yes, describe the nature and location
___________________________________________________

**Abdominal palpation:** soft, painless **YES/NO**

**Liver:** aligned with the costal arch: **YES/NO**
If no, describe in detail ____________________________________________________________

**Spleen:** not enlarged **YES/NO**
If not, describe in detail ___________________________________________________________

**Specific laboratory diagnostics:**

**Tested with PCR for SARS-CoV-2 RNA**: **YES/NO**
If yes, indicate the results ____________________________________________________

**Current therapy:** **YES/NO**
If yes, describe in detail the pharmaceuticals and daily doses________________________________

**Assessment of adverse events:** **YES/NO**
If yes, describe in detail the nature of the adverse event________________________________

**Assessment of exclusion criteria**

A healthy volunteer was excluded from the study if SARS-CoV-2 virus RNA was detected in biomaterial samples by polymerase chain reaction (PCR) for COVID-19 or if it was positive in enzyme-linked immunosorbent assay ELISA IgM and ELISA IgG: **YES/NO**

A healthy volunteer may withdraw their consent to participate in the study at any time and without giving reasons. The investigator may also decide on early discontinuation of the participation of a healthy volunteer in the study at any time if the patient's condition requires it: **YES/NO**

Main reasons for early discontinuation of the participation of a healthy volunteer in the study: **YES/NO**

Individual intolerance to the combination medication or any of its components: **YES/NO**

Failure to meet the inclusion/exclusion criteria: **YES/NO**

Refusal of a healthy volunteer to participate in the study or the withdrawal of informed consent by a healthy volunteer: **YES/NO**

Development of AEs that would prevent further prophylactic therapy with the combination medication: **YES/NO**

Emergence of a serious adverse event in a healthy volunteer: **YES/NO**

The emergence of exclusion criteria in the course of the study: **YES/NO**

Any condition of a healthy volunteer that requires, in the opinion of the investigator, the withdrawal of the patient from the study: **YES/NO**

Discontinuation of the trial by the research center, investigator, or regulatory authority: **YES/NO**

**Patient meets all the criteria of the study: YES/NO**
If not, specify the reason in detail ______________________________________________
_____________________________________________________________________________________

If the volunteer continues the trial, the date of the next visit is **_ _. _ _. _ _ _ _**

**Investigator:**_____________________________________________ ______________________
*Name Surname* *Signature*

**Date:** «____» _______________ 2020

### **Visit 3** (Treatment period)
### **Study day 14**

**Date of the visit** _ _. _ _. _ _ _ _

**Physical examination:**

**Body temperature __ __ °C**

**Swollen lymph nodes:** **YES/NO**

If yes, underline which nodes are affected: occipital, posterior cervical, parotid, anterior cervical, submandibular, sublingual, supraclavicular and subclavian, axillary, epitrochlear, inguinal, popliteal.

**Pharyngeal examination:** hyperemia **YES/NO**

**BP sitting _ _ _/_ _ _**(mm Hg)

**HR _ _ _**per min

**RR _ _** per min

**Wheezing in the lungs (dry/wet):** **YES/NO**
If yes, describe the nature and location
____________________________________________________

**Abdominal palpation:** soft, painless **YES/NO**

**Liver:** aligned with the costal arch: **YES/NO**
If no, describe in detail ____________________________________________________________

**Spleen:** not enlarged **YES/NO**
If not, describe in detail ____________________________________________________________

**Specific laboratory diagnostics:**

**Tested with PCR for SARS-CoV-2 RNA**: **YES/NO**
If yes, indicate the results _____________________________________________________

**Current therapy:** **YES/NO**
If yes, describe in detail the pharmaceuticals and daily doses________________________________

**Assessment of adverse events:** **YES/NO**
If yes, describe in detail the nature of the adverse event________________________________

**Assessment of exclusion criteria**

A healthy volunteer will be excluded from the study if SARS-CoV-2 virus RNA is detected in biomaterial samples by a polymerase chain reaction (PCR) for COVID-19 or if there is positive reactivity to enzyme-linked immunosorbent assay (ELISA) IgM and/or IgG: **YES/NO.**

A healthy volunteer may withdraw their consent to participate in the study at any time and without giving reasons. The investigator may also decide on early discontinuation of the participation of a healthy volunteer in the study at any time if the patient's condition requires it: **YES/NO**

Main reasons for early discontinuation of the participation of a healthy volunteer in the study: **YES/NO**

Individual intolerance to the combination medication or any of its components: **YES/NO**

Failure to meet the inclusion/exclusion criteria: **YES/NO**

Refusal of a healthy volunteer to participate in the study or withdrawal of informed consent by a healthy volunteer: **YES/NO**

Development of AEs that would prevent further prophylactic therapy with the combination medication: **YES/NO**

Emergence of a serious adverse event in a healthy volunteer: **YES/NO**

The emergence of exclusion criteria in the course of the study: **YES/NO**

Any condition of a healthy volunteer that requires, in the opinion of the investigator, the withdrawal of the patient from the study: **YES/NO**

Discontinuation of the trial by the research center, investigator, or regulatory authority: **YES/NO**

**Patient meets all the criteria of the study: YES/NO**

If not, specify the reason in detail ________________________________________________

If the volunteer continues the trial, the date of the next visit is **_ _. _ _. _ _ _ _**

**Investigator:**_________________________________________ ______________________
*Name Surname* *Signature*

**Date:** «____» _______________ 2020

### **Visit 4** (Follow-up period)
### Study day 21±1

**Date of the visit** _ _. _ _. _ _ _ _

**Physical examination:**

**Body temperature __ __ °C**

**Swollen lymph nodes:** **YES/NO**

If yes, underline which nodes are affected: occipital, posterior cervical, parotid, anterior cervical, submandibular, sublingual, supraclavicular and subclavian, axillary, epitrochlear, inguinal, popliteal.

**Pharyngeal examination:** hyperemia **YES/NO**

**BP sitting _ _ _/_ _ _**(mm Hg)

**HR _ _ _**per min

**RR _ _** per min

**Wheezing in the lungs (dry/wet):** **YES/NO**
If yes, describe the nature and location
_______________________________________________

**Abdominal palpation:** soft, painless **YES/NO**

**Liver:** aligned with the costal arch: **YES/NO**
If no, describe in detail _______________________________________________

**Spleen:** not enlarged **YES/NO**
If not, describe in detail _______________________________________________

**Specific laboratory diagnostics:**

**Tested with PCR for SARS-CoV-2 RNA**: **YES/NO**
If yes, indicate the results _______________________________________________

**Current therapy:** **YES/NO**
If yes, describe in detail the pharmaceuticals and daily doses______________________________

**Assessment of adverse events:** **YES/NO**
If yes, describe in detail the nature of the adverse event______________________________

**Assessment of exclusion criteria**

A healthy volunteer will be excluded from the study if SARS-CoV-2 virus RNA is detected in biomaterial samples by polymerase chain reaction (PCR) for COVID-19 or if there is positive reactivity to enzyme-linked immunosorbent assay (ELISA) IgM and/or IgG: **YES/NO.**

A healthy volunteer may withdraw their consent to participate in the study at any time and without giving reasons. The investigator may also decide on early discontinuation of the participation of a healthy volunteer in the study at any time if the patient's condition requires it: **YES/NO**

Main reasons for early discontinuation of the participation of a healthy volunteer in the study: **YES/NO**

Individual intolerance to the combination medication or any of its components: **YES/NO**

Failure to meet the inclusion/exclusion criteria: **YES/NO**

Refusal of a healthy volunteer to participate in the study or withdrawal of informed consent by a healthy volunteer: **YES/NO**

Development of AEs that prevent further prophylactic therapy with the combination medication: **YES/NO**

Emergence of a serious adverse event in a healthy volunteer: **YES/NO**

The emergence of exclusion criteria in the course of the study: **YES/NO**

Any condition of a healthy volunteer that requires, in the opinion of the investigator, the withdrawal of the patient from the study: **YES/NO**

Discontinuation of the trial by the research center, investigator, or regulatory authority: **YES/NO**

**The patient meets all the criteria of the study: YES/NO**

If not, specify the reason in detail ________________________________________________

If the volunteer continues the trial, the date of the next visit is **_ _. _ _. _ _ _ _**

**Investigator:**_________________________________________ ______________________
*Name Surname* *Signature*

**Date:** «____» _______________ 2020
